# A whole system approach to increasing children’s physical activity in a multi-ethnic UK city: a process evaluation protocol

**DOI:** 10.1101/2021.05.26.21257853

**Authors:** Jennifer Hall, Daniel D Bingham, Amanda Seims, Sufyan Abid Dogra, Jan Burkhardt, James Nobles, Jim McKenna, Maria Bryant, Sally E Barber, Andy Daly-Smith

## Abstract

**Background:** Engaging in regular physical activity requires continued complex decision-making in varied and dynamic individual, social and structural contexts. Widespread shortfalls of physical activity interventions suggests the complex underlying mechanisms of change are not yet fully understood. More insightful process evaluations are needed to design and implement more effective approaches. This paper describes the protocol for a process evaluation of the JU:MP programme, a whole systems approach to increasing physical activity in children and young people aged 5-14 years in North Bradford, UK.

**Methods:** This process evaluation, underpinned by realist philosophy, aims to understand the development and implementation of the JU:MP programme and the mechanisms by which JU:MP influences physical activity in children and young people. It also aims to explore behaviour change across wider policy, strategy and neighbourhood systems. A mixed method data collection approach will include semi-structured interview, observation, documentary analysis, surveys, and participatory evaluation methods including reflections and ripple effect mapping.

**Discussion:** Not only is this an innovative approach to process evaluation but it will also feed into iterative programme development to generate evidence-based practice and deliver practice-based evidence. This paper advances knowledge regarding the development of process evaluations for evaluating systems interventions, and emphasises the importance of process evaluation.

## 1.0 Background

### 1.1 Physical (in)activity and health inequalities

There is substantial evidence that social structural factors such as deprivation, ethnicity, gender and age influence health-related risk, health outcomes and mortality rates [1, 2, 3]. Physical activity (PA), which is positively related to health, wellbeing and academic outcomes [4, 5, 6], is also socially patterned [7]. Those who live in more deprived areas and/or are of ethnic minority populations are consistently reported to engage in lower levels of PA than less deprived and / or ethnic majority populations [8, 9, 10]. Social stratification of lifestyle behaviours, including PA, provides a partial explanation for the social inequalities of health, and can serve to perpetuate existing health inequalities [11].

### 1.2 Approaches to increasing physical activity and reducing health inequality

Increasing population levels of PA and reducing inequality is considered a public health priority [12, 13]. Until recently, PA interventions have emphasised individual-level behaviour change, which can *worsen* health inequalities, as they are often less accessible and effective for more deprived populations, due to lesser material resources and ‘leisure’ time [14]. Empirical evidence supports the proposal that behaviour is not solely the product of ‘intention’, but rather is influenced by multiple interacting forces at structural, environmental/neighbourhood, organisational, intrapersonal and individual levels [15, 16]. Interventions that target multiple ‘levels’, alter structures and processes, strengthen relationships between communities, and redistribute power resources, are more likely to increase PA behaviour and reduce inequality, than interventions that only target or that focus primarily on individual behaviour change [17, 18]. Hence, PA and health may be regarded as a “co-responsibility” of governments, individuals, families, organisations, and communities [19]. ISPAH has recently published a call to action outlining ‘eight investments that work for physical activity’. This resource advocates whole systems change across eight domains including schools, communities, travel, urban design, healthcare, workplaces, mass media, and sport and recreation [12].

### 1.3 The Bradford Local Delivery Pilot context

Responding to the need for whole systems change, Sport England has funded 12 Local Delivery Pilots (LDPs) over a 5-year period (2019-2024), to take a whole systems, place-based approach to reduce physical inactivity and health inequalities. In Bradford, 24% of residents are under the age of 16, making it the ‘youngest’ city in the UK [20]. Bradford is an ethnically diverse city - over 20% of the total district population, and over 40% of children, are of South Asian origin [21]. Bradford falls in the most deprived quintile of the Index of Multiple Deprivation, with 60% of the population living in the poorest 20% of wards in England and Wales [20]. The Bradford LDP is led by the [Born in Bradford](https://borninbradford.nhs.uk/) research programme on behalf of [Active Bradford](https://www.activebradford.com/), a partnership of organisations committed to improving physical activity within the district. Unpublished data from the Born in Bradford cohort study [22] indicates that, on average, children and young people in Bradford have lower levels of PA than the general UK population. Given the high numbers of children, and the inverse association between PA levels and age during childhood [23] the Bradford LDP, [JU:MP](https://www.activebradford.com/jump) (Join Us: Move. Play), is focused on reducing inactivity in the 27,000 children and young people aged 5-14, and their families residing in the Bradford LDP area. Further information on the programme is contained in section 2.1

The JU:MP programme is one of several system-wide interventions contributing to a major new prevention research programme called [ActEarly](https://actearly.org.uk/). The purpose of ActEarly is to identify, implement and evaluate upstream interventions within a whole system city setting. The collective aim of these multiple, system-wide interventions (including JU:MP), enacted in one locality (i.e. Bradford), is to achieve a tipping point for better life-long health and wellbeing, and to evaluate the impact of this way of working. As such, the process evaluation of JU:MP will acknowledge the broader context in which the programme is operating, including understanding which other system-wide interventions are concurrently taking place and how these interact with JU:MP to impact upon the health and wellbeing of children and young people.

### 1.4 The importance of process evaluation

Randomised controlled trials and related outcome evaluations, such as quasi-experimental controlled studies, tell us whether an intervention works in a particular setting, at a particular time, with a particular group of people. In the Bradford LDP, effectiveness studies are taking place at both population and neighbourhood levels. Better understanding of the ‘how’ and ‘why’ of the JU:MP programme, to understand the processes and interlinked contextual factors influencing change, will establish a greater appreciation of the transferability of the intervention. This is especially important for evaluating complex (systems approaches, adaptive) multi-component interventions; mechanisms influencing change are likely to be more complex, varied and dynamic [24]. Complementing an outcome evaluation with a process-oriented evaluation helps uncover processes - incorporating temporal and spatial contextual influences - influencing change [25].

Understanding how intervention (in)effectiveness arises is not the only valuable question within intervention research [26]; feasibility and acceptability are important too - alongside effectiveness, they also shape the level of embeddedness of different approaches as part of a wider whole system programme. Furthermore, process evaluations involving ongoing interaction with key stakeholders can help bridge the research-practice gap [27] and can be viewed as part of the intervention ‘system’ by providing feedback and contributing to iterative programme development [28]. A growing body of evidence - in both the health and social sciences - supports conducting process evaluations of complex interventions [e.g. 25, 29]. However, few PA evaluations have captured the complexity of behaviour change systems [30]. This paper describes the protocol for a process evaluation of the development, implementation and evaluation of the JU:MP programme.

## 2.0 Methods

This paper focuses on the process evaluation of the JU:MP programme approach. Reporting is guided by the RAMSES II reporting standards for realist evaluations; see additional file 1. The process evaluation will be conducted alongside a complementary effectiveness evaluation and findings from across the broader evaluation will be integrated to advance knowledge production [31].

### 2.1 Intervention: The JU:MP Programme

The underlying themes, framework (tool, settings and principles) and theory of change for the JU:MP programme were developed in 2018 based on community consultation and priority setting workshops, data from the Born in Bradford research programme [22, 32, 33], international peer-reviewed evidence [34, 35] and the socio-ecological model [36]. Subsequently the first iteration of the JU:MP implementation plan was designed, with projects aligned to the programme themes and content related to the theory of change. During 2019-2020 a test-and-learn phase was undertaken, ‘pathfinder’. In 2021, based on the experiences from the ‘pathfinder’ phase a second version of the implementation plan was drawn up. This included the creation of the JUMP model depicting 15 workstreams which will be taken forward into the delivery of the ‘accelerator phase’ (2021-2024). JU:MP has been designed for continuous improvement, based on process evaluation and learning. The description here reflects JU:MP as we transition from the pathfinder phase (the initial small scale test and learn period over 2019-2021) to the accelerator phase (the roll-out of the developed programme across the LDP over 2021-2024); see Figure 1 for a timeline illustrating key milestones. JU:MP is seen as a whole system approach; the theory of change outlines five themes (family, community, organisations, environment, and policy and strategy) through which JU:MP will ‘act’ to increase PA in children aged 5-14 years, and subsequently improve wider health and social outcomes (see Figure 2). While the underlying theory of change incorporates multiple ‘mechanisms’, it is recognised that JU:MP is both a system-based intervention and is being implemented within a complex social system, where the process of change in reality will be complex, messy and nonlinear. As such, the theory of change does not provide an exhaustive list of practice-based mechanisms. Four guiding principles underpin the JU:MP approach: i) tailored approaches to change and to link levels within a whole system; ii) community involvement at every step of the process; iii) engaged, active leaders and partners; and iv) evidence- and insight-led. The implementation plan includes 15 interacting work streams which cut across the five JU:MP themes. There are six overarching work streams that are delivered across the whole LDP area, and nine that are developed and delivered at a neighbourhood level (see Figure 3).

**Figure 1.**
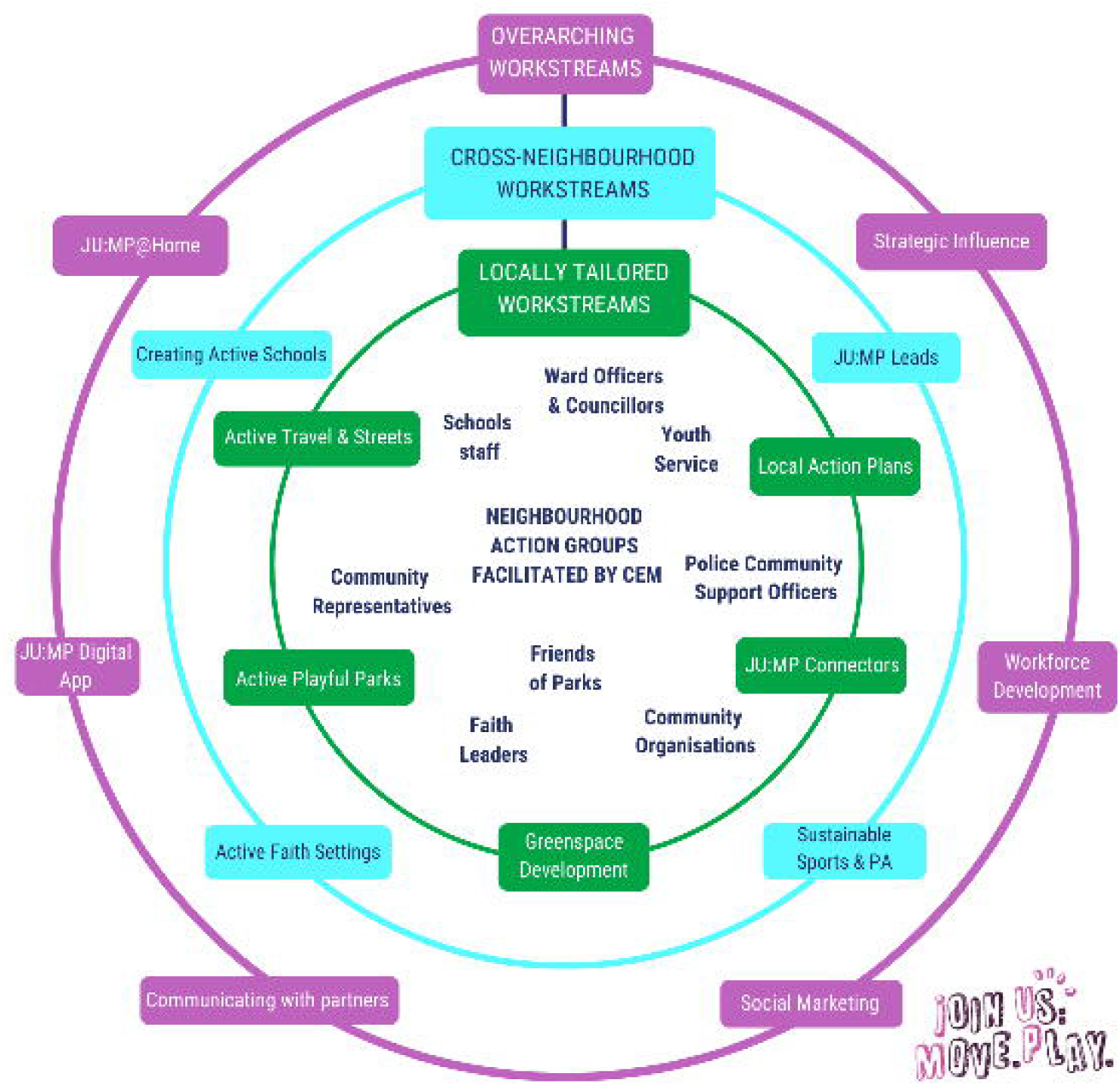
JU:MP programme timeline (key milestones)

**Figure 2.**
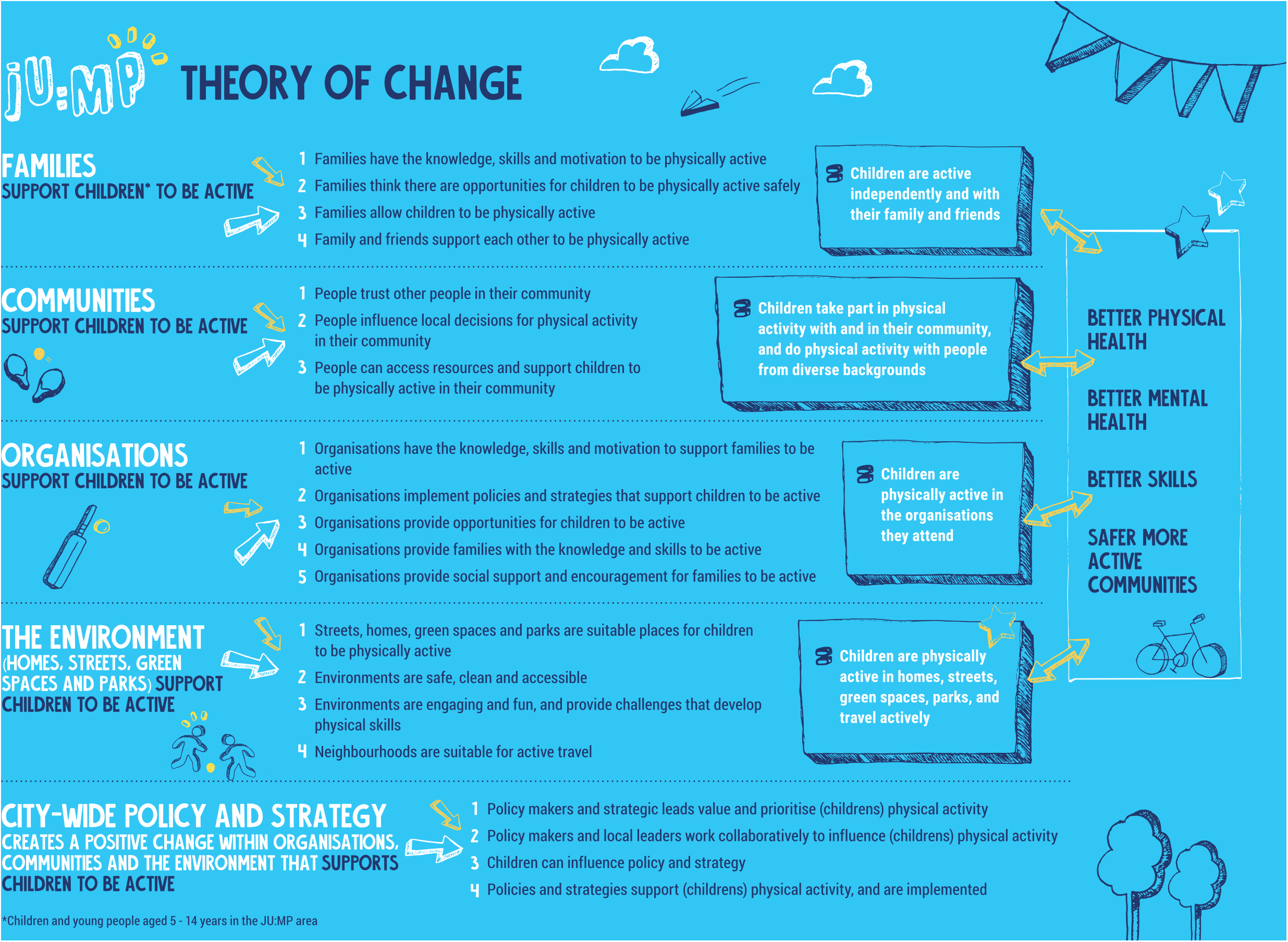
JU:MP theory of change

**Figure 3.**
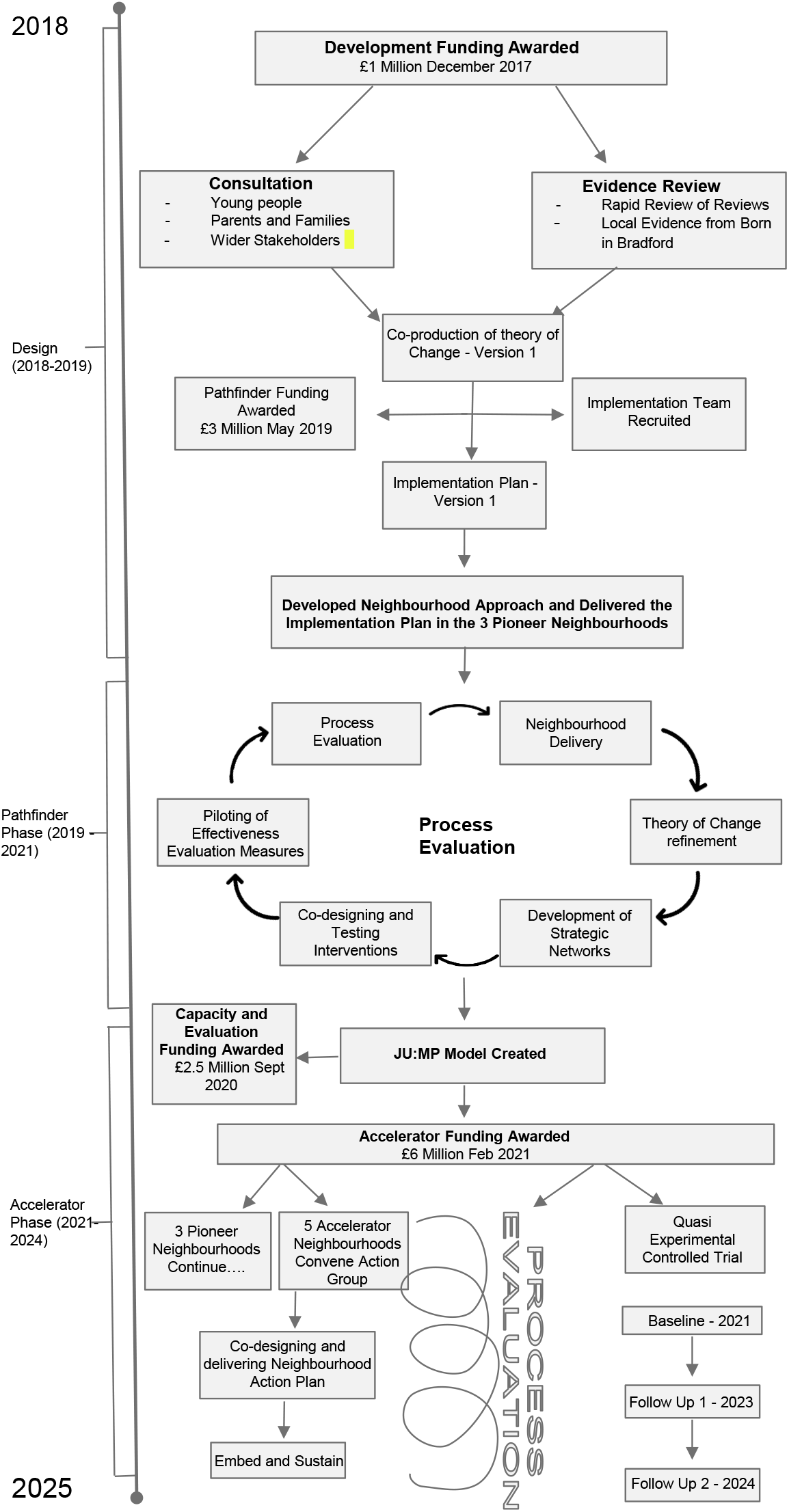
The JUMP programme model

JU:MP is being implemented within eight distinct geographic ‘neighbourhoods’ within the Bradford LDP area; see additional file 2 for a map of the LDP neighbourhoods. Neighbourhood boundaries were based on areas having an area of green space with potential for development, at least 4-5 primary schools, and an active community organisation. This hyper-local scale of whole systems delivery aims to foster genuine collaborative working and building strong sustainable relationships. Using an asset-based community development approach, JU:MP facilitates the development of an action group within each neighbourhood, including key organisational partners, community members, and families. To allow the programme to meet local needs and facilitate longer-term behaviour change, the action group is jointly responsible for (1) co-producing local action plans and green space developments (approximately three months), (2) collectively delivering the local action plans, with members contributing to delivering separate work streams (e.g. school stakeholders deliver Creating Active Schools) (approximately one year) and (3) the ‘embed and sustain’ phase during which time JU:MP facilitation is lessened (approximately one year).

Initially, the neighbourhood approach was operationalised within three ‘Pioneer Neighbourhoods’ (pathfinder phase - 2019-2021) to undertake a test and learn process. Subsequently, the programme will be delivered in the five remaining neighbourhoods (2021-2024), to cover the whole LDP area. The accelerator phase neighbourhoods are further broken down into those that are directly facilitated by the JU:MP team, as in the pathfinder phase (n = 3), and those whose delivery will be externally commissioned (n = 2). The programme model is illustrated in Figure 3. Additional file 3 offers a more detailed description of each work stream.

### 2.2 Process evaluation theory: realism, systems thinking, complexity science

A realist philosophy underpins this process evaluation. Realism posits that an objective reality exists, but that knowledge is ‘value-laden’ and as such we can only understand reality from within a particular discourse [37]. Realism holds that reality exists in an open-system, meaning that attention in programme development and evaluation perspective focuses on how context and mechanisms interact to influence outcomes [38]. Process evaluation is typically understood as “the evaluation of a process of change that an intervention attempts to bring about in order, at least in principle, to explain how outcomes are reached” [29].

Underpinned by realist principles, the role of context is prioritised in establishing intervention (in)effectiveness [29]. Examining context implies focusing on social processes to establish an understanding of how different notions of intervention feasibility, acceptability and effectiveness can be framed. Another part of realist evaluation allows the development and / or refinement of theories relating to mechanisms of change, focusing on context-mechanism-outcome configurations. This supports the iterative development of programme logic models and theories of change [39]. However, a realist approach acknowledges that people attach meaning to experiences, and meanings are implicated within causal processes [37]; behaviour therefore cannot be fully explained, as people are conscious beings that act back on the structures and processes of social life [40].

Within the complex intervention evaluation field, recent calls to embed complexity science and systems principles within process evaluation design reflect a move towards understanding how interventions are part of complex adaptive systems [41, 42, 24]. Realist methodology is consistent with systems thinking and complexity science [41]. They share a mutual belief that wider contexts are inherent within change mechanisms [39]. Yet, systems thinking necessitates taking a holistic view to examine how systems (including interventions) influence behavioural change, rather than viewing interventions in isolation. Further, complexity science is concerned with how interactions between different system elements (including interventions) create change, focusing on concepts including dynamism, nonlinearity, adaptation, feedback loops, and co-evolution [41, 42].

Realism, systems thinking, and complexity science have shaped the development of the aims, study design, data collection, and analysis of the JU:MP process evaluation. Predominantly qualitative methods have been adopted here, using a longitudinal design, to establish a fuller understanding of intervention acceptability and effectiveness, and to capture how acceptability and effectiveness change as systems evolve e.g. generate feedback loops [39, 42].

### 2.3 Aims, objectives and approach

The overarching aim of the process evaluation is to understand the programme implementation and the mechanisms through which JU:MP influences behaviour change across the neighbourhood, and wider policy and strategy systems that it is seeking to influence. The evaluation also facilitates dynamic system change via informing the refinement of the programme and associated theory of change. To address these aims, and in accordance with the JU:MP delivery approach, the process evaluation includes three distinct but interrelated packages of work: (1) a strategic-level evaluation, (2) a neighbourhood-level evaluation, and (3) an end-user evaluation. Table 1 provides an overview of the scope and objectives of each process evaluation work package.

**Table 1.** The scope and objectives of the strategic, neighbourhood and end user-level process evaluation work packages

|  | **Strategic-level** | **Neighbourhood-level** | **End-user level** |
| --- | --- | --- | --- |
| **Scope**<br>(aims, stakeholders) | To understand the views and actions of JU:MP strategic-level stakeholders, including the core JU:MP team and executive board, stakeholders commissioned to lead on the strategic delivery of work streams, and city-wide strategic partners such as the Living Well programme strategic leads | To understand the views and actions of stakeholders involved in developing and / or implementing JU:MP within a JU:MP neighbourhood (e.g. voluntary organisation stakeholders, school leads, councillors, faith setting leads, friends of groups, families) | To understand the views and actions of the 'end user' recipients of JU:MP, i.e. children and young people and their families living in North Bradford |
| **Objective**<br>(documentation) | To document the strategic-level design, delivery and evaluation processes of the JUMP programme, including: individual work streams and evaluation packages and interactions | To document JU:MP programme neighbourhood level design and delivery processes, including: (a) the community engagement and co-production process and (b) the design and implementation of the overarching action plan and specific interventions | n/a |
| **Objective**<br>(feasibility and acceptability) | To examine the feasibility and acceptability of the strategic level design, delivery and evaluation of the JU:MP programme, by understanding the barriers, facilitators and contextual factors influencing design, delivery and evaluation, including: (a) Individual work streams and evaluation packages and interactions and (b) strategic influencing across the wider system | To examine the feasibility and acceptability of the neighbourhood level design and implementation of the JU:MP programme, by understanding the barriers, facilitators and contextual factors influencing design and delivery, including: (a) examining the feasibility and acceptability of the neighbourhood co-production approach and (b) examining the feasibility and acceptability of delivering the overarching plan and specific interventions | To examine the experience of children and families receiving JU:MP, including understanding the JU:MP 'journey' and acceptability of JU:MP for different users |
| **Objective**<br>(impact) | (a) To understand the impact of JU:MP across the whole system including unintended consequences, and developing an understanding of change mechanisms (what works, for whom, and in what context) from the perspective of strategic-level stakeholders<br>(b) To understand the impact of JU:MP on strategic-level stakeholders<br>(c) to understand the impact of JU:MP on city-wide policy and strategic working around physical activity | (a) to understand the impact of JU:MP across and beyond the neighbourhood system including unintended consequences, and developing an understanding of change mechanisms (what works, for whom, and in what context) from the perspective of neighbourhood-level stakeholders<br>(b) to understand the impact of JU:MP, on neighbourhood-level stakeholders | To understand the impact of JU:MP including unintended consequences, and change mechanisms |

#### 2.3.1 Strategic-level process evaluation study design

A longitudinal mixed methods design is being adopted. The study received ethical approval from Leeds Beckett University in March 2020 (ref: 69870), and will run until programme delivery ceases in 2024. The overarching objectives of the strategic-level process evaluation are to document, and understand the feasibility, acceptability and impact of the strategic-level development, delivery and evaluation of JU:MP. This includes a focus on the 15 JU:MP programme work streams (see Figure 2), and the effectiveness, process and individual project evaluations, including interaction, synergy and tension between these different JU:MP system elements.

Stakeholders involved in the development and delivery of JU:MP at a strategic-level, i.e. beyond individual neighbourhoods, will be invited to participate. Data collection methods include surveys, semi-structured interviews, participant observations, and reflections, which will all be implemented at multiple time points throughout programme delivery; see section 2.6 for further detail. The evaluation will iteratively refine as priorities surface; for example, we have recently incorporated a sub-study to provide a ‘deep dive’ into the strategic-influencing work of JU:MP to examine the wider intended and unintended impacts of city-wide policy and strategic working related to PA, following the addition of policy and strategy as a theme within the theory of change.

#### 2.3.2 Neighbourhood-level process evaluation study design

A longitudinal, mixed-methods case study design is being adopted, with individual neighbourhoods being classified as ‘cases’. This study received ethical approval from the University of Bradford in November 2020 (ref: E838) and will be implemented during the ‘delivery’ phase within each JU:MP neighbourhood, which lasts approximately three years. Section 2.1 provides detail on the neighbourhood delivery approach.

A minimum data-set will be collected from each neighbourhood, with additional data collection occurring within selected ‘deep dive’ neighbourhoods. These neighbourhoods will include one from the pioneer neighbourhood phase (with the primary aim of piloting and refining the data collection techniques, and to inform programme design and delivery), and the three accelerator phase neighbourhoods that are directly facilitated by the JU:MP team, in line with the neighbourhoods that are included within the neighbourhood control trial that forms part of the effectiveness evaluation of JU:MP. Aligning the ‘deep dive’ neighbourhoods with those included in the control trial will generate greater understanding and explanation of control trial findings; the trial will provide evidence of JU:MP effectiveness within the neighbourhood. The process evaluation will help explain what worked, why, when, for whom, and within what context.

Amendments to the evaluation protocol will be made following piloting and prior to implementing the study within the ‘accelerator phase’ neighbourhoods. Minimum-data data collection methods include surveys and documentary analysis, and additional methods employed in ‘deep dive’ neighbourhoods include extra surveys, process observations, semi-structured interviews, and participatory evaluation methods; see section 2.6 for further detail.

#### 2.3.3 End user-level process evaluation study design

The end-user process evaluation will examine the experiences and impact of JU:MP amongst children and families. This will feature focus groups with children and parents/guardians from across the accelerator direct delivery neighbourhoods, approximately 12 months and 24 months following JU:MP commencement. Additionally, in-depth longitudinal research will be conducted with approximately four local families. Citizen science methods will be adopted, which involves members of the public (non-scientists) collecting and analysing data, in collaboration with researchers [38], to foster community engagement.

Multiple and innovative methods of data collection will be employed, which *could* include written or video diaries, or photo-elicitation techniques, walk-and-talk interviews, but crucially, the families will be engaged in developing the research approach, collecting and analysing their own data, and making recommendations for future practice. A PhD studentship, jointly funded by Sport England (as part of the programme funding) and the University of Bradford, will develop and conduct this work, commencing in 2021. It is preemptive to give close detail of methods and analysis for an area of work that is still emerging.

### 2.4 Theories and models utilised within the process evaluation

Various existing theories / models / frameworks underpin the development, delivery and evaluation of the JU:MP programme. Herein, we focus on theories that are used directly, or indirectly as sensitising concepts, within the process evaluation of JU:MP, including in the development of topic guides and surveys, and analysis frameworks.

1. Consolidated framework for implementation research (CFIR) [44] - The CFIR was developed by synthesising implementation constructs from across 20 implementation sources and multiple scientific disciplines [44], and is a comprehensive framework designed to examine intervention implementation [45]. Five major domains comprise the CFIR: intervention characteristics, inner setting, outer setting, characteristics of individuals involved in implementation, and the implementation process [44]. The CFIR is being used as a sensitising framework within the process evaluation to understand the feasibility of implementing the JU:MP programme.
2. Capability, opportunity, motivation-behaviour (COM-B) [46] and the Theoretical Domains Framework (TDF) [47] - The COM-B model provides a comprehensive and evidence-based model for understanding human behaviour and behaviour change. The model proposes that behaviour is influenced by capability (physical, psychological), opportunity (physical, social) and motivation (reflective, automatic), and that all three must be present for a behaviour to occur [46]. The TDF consists of 14 ‘domains’ of influence on behaviour, developed by synthesising 33 theories of behaviour and behaviour change [47]. The Domains align to COM-B categories and can be used to develop and implement interventions and to inform understanding of the barriers and facilitators influencing behaviour change.
3. JU:MP programme theory of change - The programme theory (see section 2.1 and Figure 1) is implicated in the evaluation of JU:MP; it will be utilised to understand impact and mechanisms of impact as well as being iteratively refined as programme delivery and evaluation progress.

### 2.5 Sampling and recruitment

The proposed sample for the strategic-level study includes stakeholders who are part of the strategic leadership of the JU:MP programme. This includes all members of the core JU:MP research and implementation teams, stakeholders commissioned to lead on the strategic delivery of one of the 15 work streams across the LDP, JU:MP executive board members and members of the established strategic development working group for integrating physical activity in policy and strategy across the district. The sample size will be based on the number of individuals that meet the inclusion criteria, which is expected to be around 100. The proposed sample for the neighbourhood-level study includes stakeholders who are involved in designing and delivering JU:MP within one (or more) of the participating neighbourhoods, as part of the neighbourhood action group, including for example, JU:MP connectors, Islamic Religious Setting stakeholders, and children and families; see section 2.1. The sample size is based on the expected number of individuals (20) that will form the action groups within each neighbourhood, meaning there will be around 160 participants in total. As detailed in section 2.6, not all participants will take part in all aspects of data collection, for example interviews will only be conducted with approximately 20 individuals at each time point in both the strategic and neighbourhood-level studies.

All potential participants across both the strategic and neighbourhood level studies will be engaged in the design and delivery of JU:MP, and as such will already be known and identifiable to the research team, via the implementation team. Potential participants will be given an information sheet for the research, and informed consent will be obtained prior to data collection commencing. Data collection will take place at multiple time-points over a significant time-period (up to for years). At each data collection ‘point’, participants will be verbally reminded that they are taking part in the study and what it involves, and will be given a verbal reminder to let the researcher know at any time if they wish to withdraw their consent to participate.

### 2.6 Process evaluation data collection methods

This section provides a rationale for and description of each data collection method that is being utilised within the JU:MP process evaluation. Table 2 provides a map of when and where each method is being utilised as part of the strategic and neighbourhood evaluation work packages.

**Table 2.** Data collection methods for the strategic and neighbourhood process evaluation.

| **Data collection method** | **Strategic level process evaluation** | **Neighbourhood level process evaluation** |
| --- | --- | --- |
| **Surveys** | Participant characteristics survey: upon recruitment (All recruited participants)<br>Influences on behaviour survey : every 6 months (All recruited participants) | Participant characteristics survey: upon recruitment (All participants, all neighbourhoods)<br>Influences on behaviour survey: baseline, 6-months, 12-months and 24 months (All participants, all neighbourhoods)<br>Network mapping survey: baseline, 6-months, 12-months and 24 months (All recruited participants, all neighbourhoods) |
| **Process observations** | Process observations of meetings including:<br>Implementation team meetings: one in every four (attended by core team members such as the programme director, community engagement managers and communications officer)<br>Other key strategic meetings identified in collaboration with the implementation team | Process observations of action group workshops: every workshop, approximately once every six weeks (deep-dive neighbourhoods only) |
| **Documentary analysis** | Key documents for each work stream collated every 6 months (including service agreements, project plans and evaluations) | Action group workshop notes (All neighbourhoods)<br>Neighbourhood action plans (All neighbourhoods) |
| **Semi-structured interviews** | Interviews with around 20 strategic stakeholders every 6 months (including members of the core team and one strategic lead for each workstream at each time point)<br>Interviews with around six additional wider stakeholders every 12 months (three members of the executive board and three members of the strategic development) | Interviews with around 20 neighbourhood stakeholders at 6 and 18 months (including key delivery stakeholders such as JU:MP connector, Islamic Religious Setting lead, school lead etc. from across deep-dive neighbourhoods only)<br>Interviews with around two commissioned organisation stakeholders at 6 and 18 months (commissioned neighbourhoods) |
| **Reflections** | Group reflections embed into key meetings:<br>Weekly implementation team meetings: one in every four (attended by core team members such as the programme director, community engagement managers and comms officer)<br>Weekly research team meetings: one in every four (attended by core team members such as the research directors and research fellows) | - |
| **REM** | REM workshops embedded into strategic development group meetings: every 6 months | REM workshops embedded into action group meetings: every 6 months (all neighbourhoods) |

#### 2.6.1 Surveys

a. Personal characteristics survey - a short survey related to the participants’ personal characteristics, including gender, date of birth, home postcode, ethnicity, highest qualification, employer, job role, and JU:MP role(s). This survey will enable characterisation of the sample and will aid in contextualising and interpreting qualitative data.
b. Influences on behaviour survey - the survey has been developed to assess factors influencing participants’ roles in supporting the design and delivery of JU:MP. The survey is an adapted version of a validated 6-item COM-B questionnaire [48]; see additional file 4 for a copy of the survey. Draft surveys were piloted with members of the core team, and refined based on feedback. The survey will permit the identification of determinants of behaviour [49], which will highlight areas for intervention to increase the capability, opportunity and / or motivation of stakeholders to influence change and to support children to increase physical activity. Repeating the survey at baseline, 6 months, 12 months and 24 months will permit an understanding of how different influences change over time. Further exploration during interviews for some participants will aid in understanding the reasons for these changes.
c. Stakeholder mapping survey - this survey has been developed to facilitate a social network analysis [50]; connections between stakeholders will be mapped to understand the impact of JU:MP on relationships between parties within neighbourhood networks. Published guidance on social network analysis [51] and input from network analysis specialists informed the initial development of the survey. The survey is being refined following piloting with pioneer neighbourhood stakeholders. Repeating the survey every six months will permit an understanding of how relationships develop and change over the course of the JU:MP programme. See additional file 5 for a copy of the stakeholder mapping survey.
d. Feedback forms - feedback forms will be administered following neighbourhood action group workshops to examine participants’ thoughts and feelings about the workshop content and process, and to understand the emerging impact of the work. The content of the forms may be adapted depending on the workshop context, however questions will typically include “What did you find most useful about the workshop?”, “What did you find least useful about the workshop?, How could it be improved?” and “What is the most significant output of JU:MP so far?”

#### 2.6.2 Semi-structured interviews

Semi-structured interviews provide an opportunity for in-depth reflection on the design and delivery of JU:MP, including documenting and reflecting on progress, activity, decisions, perceptions, and challenges [52]. Understanding these processes is important for evaluation, as it helps us to understand the factors influencing whether or not the programme is successful in achieving its outcomes. The interviews will explore the capability, opportunity and motivation of the participants to support the JU:MP programme and will be tailored to their specific role within JU:MP. The opportunity to reflect on involvement via an in-depth interview can also have a positive influence on programme design and delivery via facilitating a process of continuous learning [53]. Interview guides are theoretically informed; they draw on implementation theory (CFIR), behavioural theory (COM-B and TDF), and the JU:MP theory of change. However, interview guides will be refined on an iterative basis based on project developments and prior data collected via other methods, for example, observations (see section 2.6.3).

#### 2.6.3 Participant observation

Observation offers a direct view of behaviour, capturing events as they occur in their natural setting [54]. Qualitative observations, completed by a researcher, provide an independent record of activities, including developing an understanding of context, behaviours and interactions, allowing reflection on these activities [29]. Key meetings (table 2) will be observed by a researcher. Informed by Spradley [55] and aligned with the theories and frameworks underpinning programme design and evaluation (systems thinking, JU:MP ToC, COM-B, CFIR), an observation summary sheet has been prepared to guide this collection of observational data. This guidance provides common areas of focus across observational records; additional file 6. During the observation the researcher will record a ‘condensed account’ of the event, which will then be utilised as an aide-memoire to develop an expanded account. These expanded accounts will be included in data analysis.

#### 2.6.4 Documentary analysis

Key programme documents can provide insight into the design and delivery of programmes, including information on decisions made / agreed actions and why, and implementation challenges. Meeting and workshop notes and neighbourhood action plans will be included in qualitative analyses (table 2). Additionally, key documents such as service agreements and project plans will be requested from stakeholders prior to interviews, to aid the interview process e.g. discussing how and why plans were delivered as intended or amended.

#### 2.6.5 Participatory evaluation methods

a. Reflections - Regularly reflecting on programme activity is important as it allows us to document progress, activity, decisions, and challenges, as they are occurring. Reflective practice can also have a positive influence on neighbourhood design and delivery via facilitating a process of continuous learning [53]. Short reflection activities are being embedded into key JU:MP meetings, with attendees being given 60-90 seconds each to share a key learning (what happened, why and how, context, future planning) Reflections are captured as part of documentary analysis; see section 2.6.4.
b. Ripple effects mapping (REM) - This participatory method takes a qualitative, collaborative approach to understanding wider programme impacts. Unlike traditional impact evaluation methods, which tend to focus on a small number of pre-specified outcomes, REM is designed to uncover a wider range of intended and unintended impacts stemming from a programme [56]. This may be particularly important in whole systems programmes and where interventions are co-produced, flexible and emerging. The method involves holding researcher-facilitated workshops with the participants (approximately 12 per workshop) involved in developing and delivering (an aspect of) the programme, to create a visual output of impacts [57, 58]. The workshops involve four steps: team-based conversations, mapping activities and impacts, reflecting further on impacts, and identifying the most and least significant changes. The workshops can be repeated over time to understand impact pathways and timelines [59]. Previous research has documented that participating in the mapping process and realising the range of impacts can be motivating to stakeholders and encourage further action [56, 60]. Within the process evaluation, this method will be used to examine the impact of the strategic influencing work, and the neighbourhood programmes involved in deep-dive evaluation.

### 2.7 Data analysis

#### 2.7.1. Qualitative data analysis

Qualitative data including semi-structured interview data, reflections, key documents including meeting notes, and process observation summaries will be analysed using a framework approach [61]. Framework analysis is a type of thematic analysis aimed at providing descriptive and/or explanatory findings clustered around themes. Uniquely, framework analysis features using a matrix to systematically reduce the data. The key steps involved include (1) familiarisation, (2) identifying a thematic framework, (3) indexing (applying the thematic framework to the data set), (4) charting (entering data into framework matrices), and (5) mapping and interpretation [56].

A framework approach was selected for a number of reasons. First, the matrix permits multiple comparisons, including between interventions, subjects, data sources and time points [62]. This is particularly important for evaluating the JU:MP programme to allow findings to be examined both within and across different interventions within the system, and over time. Second, a framework aids in consolidating data across themes, identifying broad ranging data - discussing different JU:MP interventions, via different methods, at different time points. The framework also allows the isolation of specific data from different interventions, neighbourhoods, stakeholders etc. to be analysed separately, if required. Third, the indexing and charting process allows all members of a multidisciplinary team to engage with the analysis (e.g. of a particular theme) without needing to read and code all the data [63]. Finally, the approach is suited to prolonged data collection, allowing analysis to occur alongside data collection. This allows findings to inform iterative programme development, and ‘chunks’ analysis across the timeframe of the programme.

An initial framework was developed based on theory underpinning the evaluation, the JU:MP programme structure (deductive), and inductive coding of a small number of initial interview transcripts. Over the course of the pathfinder phase, the framework was iteratively refined based on coding of data, and the development of the programme. The refined framework includes separate themes for the different work streams and evaluation work packages, as well as themes for the overarching programme development, delivery and evaluation (additional file 7). Following coding using NVivo 12.0, framework matrices will facilitate the interpretation of data and the construction of themes. Miro will be used to visually illustrate the REM maps, while qualitative content analysis, using NVivo 12.0, will analyse the data within the REM outputs. This type of analysis will identify data patterns and quantify emerging aspects of programme outputs.

#### 2.7.2 Quantitative data analysis

Data from the participant characteristics survey and influences on behaviour survey will be summarised using descriptive statistics. Univariate statistical tests will be used to examine differences between different groups of participants, and general linear models will explore any differences in influences on behaviour over time. The network mapping survey will be analysed and illustrated using social network analysis software.

#### 2.7.3 Mixed-method integration and evidence-practice feedback loops

Following initial analysis as described in sections 2.7.1 and 2.7.2 the data will be integrated to establish context-mechanism-outcome configurations, to understand what works, when, how, and in what context [39]. Ongoing analysis will inform the refinement of the programme and associated theory of change. To facilitate this process, bi-annual process learning workshops will take place with the core JU:MP research and implementation team. Emerging findings will be presented and, using Driscoll’s learning cycle [64], the team will consider the implications of the findings and agree on changes to the programme design, how the programme is delivered, and/or how the team work (together). These changes will then be captured in the ongoing evaluation as part of workshop notes and interviews, thus completing the cycle.

## 3.0 Discussion

This paper outlines the protocol for a process evaluation of JU:MP, the Bradford LDP, a whole systems programme for increasing PA in children and young people aged 5 - 14. The aim of the process evaluation is to understand the mechanisms through which JU:MP influences PA, and to examine behaviour change across the wider policy and strategy and neighbourhood systems. The evaluation also facilitates dynamic system change via informing the refinement of the programme and associated theory of change. To address these aims, evaluations are taking place at the strategic, neighbourhood, and end-user level. Mixed methods are being employed including surveys, interviews, and process observations, and participatory methods including reflections and ripple effect mapping.

Publishing a protocol for the process evaluation of the JU:MP programme is intended to both highlight the importance of process evaluations in evaluating complex interventions, and to add to the process evaluation methodology literature. While protocols of process evaluations of PA programmes are now appearing in the literature [e.g. 65, 66, 67], typically they describe protocols for process evaluating individual interventions. In this context, our plan is a rare example that addresses a whole system programme incorporating multiple interventions [63, 64]. A key strength is that our approach remains flexible to iterative development of the programme; it is not constrained by requiring substantial ethical amendments, nor by pre-specified outcomes [56], while still ensuring that the protocol is clear, detailed and has fixed parameters for transparency and replicability purposes. At the same time, while the in-built processes can ensure evaluation is delivered as planned, they can also record any required adaptations. This paper also advances the literature by outlining a novel approach to evaluating a whole system programme, incorporating innovative participatory methods that permit iterative refinement of the programme alongside implementation [28, 27].

Given the time often required to conduct robust qualitative work, a challenge here is ensuring that the process evaluation findings remain ‘relevant’ as the JU:MP programme progresses and evolves in an agile way It is, therefore, important to ensure that the findings are fed back in a timely manner and in an appropriate format to allow the team to ‘step back’ and engage in systematic planning. Whilst the evaluation outlined in this paper is resource-intensive, it is set up to generate a deep and rich understanding of the processes underpinning programme design, implementation and impact, and thus will be invaluable in supporting other communities to apply a similar approach and / or to learn from things that have not delivered expected successes.

An embedded research team is critical for the development of research-practice partnerships, which facilitates evidence-based practice, and the development of practice-based evidence through the JU:MP programme [70]. However, a limitation of this approach is that it reduces the impartiality of the research team and thus the independence of the evaluation [71]. Successfully negotiating a suitable balance of involvement with, and detachment from, the JU:MP programme is critical to the success of the process evaluation [72]. For example, it was imperative that the research team worked alongside the programme team to develop a protocol that aligns with and meets the needs of the programme, and involvement is also required to produce detailed and in-depth observational records that reflect participant experiences. Detachment is also required throughout the research process, for example when analysing data, to ensure that the analysis is reality-congruent and theoretically informed, rather than a reflection of the researcher’s experiences within the setting.

The process evaluation outlined within this paper forms part of a wider evaluation approach, which includes an effectiveness evaluation (neighbourhood control trial, and a before and after evaluation using the Born in Bradford birth cohort). Process evaluations can be complementary to outcome evaluations, as the approaches produce different types of knowledge about a phenomenon that can be combined to further advance knowledge [24, 73]. The JU:MP programme evaluation provides an opportunity for mixed methods evidence synthesis, combining the advantages of controlled trials in estimating intervention effects, with an in-depth understanding of participants’ experiences and the mechanisms underpinning change [39, 25]. However, in doing so, it is important that the inherent value of process evaluation is appreciated, beyond facilitating interpretation of trial findings, to avoid perpetuation of the paradigmatic hegemony existent within intervention evaluation research [74].

## 4.0 Conclusion

Despite significant efforts to address children’s physical inactivity by researchers, practitioners and policy makers, physical activity levels are socially stratified, which can serve to perpetuate health inequalities [7]. Sport England has invested significant funds in 12 LDPs to increase PA and reduce inequalities through taking a place-based, whole systems approach. Methodologically rigorous, high quality research is required to examine what works, why, for whom, and in what context, to understand both the potential of whole system approaches for increasing children’s PA, and whether and how they can be replicated in other geographical contexts. The process evaluation of the Bradford LDP aims to address this.

## Data Availability

The dataset that will be generated and analysed during the current study are not publicly available to reserve the anonymity of research participants.

## 5.0 Abbreviations

BCW: Behaviour Change Wheel
COM-B: Capability, Opportunity, Motivation-Behaviour
DD: Direct Delivery
JU:MP: Join Us: Move. Play
LDP: Local Delivery Pilot
REM: Ripple Effects Mapping

## 6.0 Declarations

### Ethics approval and consent to participate

Strategic-level process evaluation, Leeds Beckett University (69870). Neighbourhood-level process evaluation, Bradford University (E838).

### Consent for publication

Not applicable.

### Authors’ contributions

This study was conceived and designed by JH and ADS, with input and feedback from all authors. The manuscript was initially drafted by JH. Subsequent drafts were commented on by all authors and revisions were made by JH. All authors have approved submission.

## Acknowledgements

We are grateful for the funding provided by Sport England to implement and evaluate the programme. The authors wish to thank the JU:MP team and wider stakeholders, including colleagues within Born in Bradford and ActEarly, including Dr Laura Sheard and Dr Bridget Lockyer, for their input in developing the process evaluation and their ongoing engagement with and openness to the evaluation process. The authors also wish to thank Aira Armanaviciute, Catherine Sadler, Dionysia Markesini, Emma Young, Isobel Steward, Lisa Ballantine, Simona Kent-Saisch, and Zubeda Khatoon for supporting the testing and refinement of the framework for analysing the qualitative data.

## 10.0 Additional files

Additional file 1. RAMSES reporting checklist. A table outlining how and where the protocol conforms to the RAMSES II reporting standards for realist evaluations. File extension: docx

Additional file 2. JU:MP neighbourhood map. A geographical map of the JU:MP neighbourhood boundaries within the LDP area. File extension: png

Additional file 3. JU:MP programme plan. A table providing a description of each of the 15 JU:MP work streams. File extension: docx

Additional file 4. Influences on behaviour questionnaire. A survey, adapted from Keyworth et al. [42], used to understand the capability, opportunity, motivation and behaviour of neighbourhood-level stakeholders related to supporting children to be physically active. File extension: doc

Additional file 5. Network mapping survey. A survey to map connections and relationships between different stakeholders and organisations within JU:MP neighbourhoods. File extension: pdf

Additional file 6. Observational framework. A framework designed to guide the collection and recording of observational data. File extension: doc

Additional file 7. JU:MP thematic framework. The framework used to index data as part of the qualitative framework analysis. File extension: doc

