## Additional file 4 for "A whole system approach to increasing children’s physical activity in a multi-ethnic UK city: a process evaluation protocol"

Your role in supporting children and their families to be physically active

This part of the questionnaire aims to understand the extent to which you feel you have the skills, motivation and opportunity to support children and young people (aged 5-14 years) and their families to be physically active.

**When we talk about activities you engage in to support children and young people to be physically active, we are referring to anything that helps children and their families to be active, whether directly or indirectly.** This could include activities you undertake as part of JU:MP, for example supporting the development and delivery of the local action plan for increasing physical activity, integrating physical activity into policy documents, making improvements to parks and green spaces, and speaking to children and families about what would help them to be more active.

It is not a test of your knowledge but a way for us to find out some  of the barriers you face in completing your JU:MP role and supporting children and families to be physically active, so please answer  these questions as honestly as you can. All responses provided will be anonymised and at no point will your individual responses be able to be identified in any research outputs.

**Please rate your agreement with the following statements**

1. I support children and their families in [neighbourhood] to be physically active at every opportunity

0	1	2	3	4	5	6	7	8	9	10

☐	☐	☐	☐	☐	☐	☐	☐	☐	☐	☐

2. My efforts to support children and their families in [neighbourhood] to be physically active are effective

0	1	2	3	4	5	6	7	8	9	10

☐	☐	☐	☐	☐	☐	☐	☐	☐	☐	☐

3. I have the PHYSICAL opportunity to support children and their families in [neighbourhood] to be physically active

**What is PHYSICAL opportunity?**

The environment provides the opportunity to engage in supporting children and their families to be active.

(e.g. enough time, appropriate funding and resources, communication channels)

0	1	2	3	4	5	6	7	8	9	10

☐	☐	☐	☐	☐	☐	☐	☐	☐	☐	☐

4. I have the SOCIAL opportunity to support children and their families in [neighbourhood] to be physically active.

**What is SOCIAL opportunity?**

Interpersonal influences, social cues and cultural norms provide the opportunity to support children and their families to be physically active

(e.g., support from colleagues, community residents and families, aligns with professional role and responsibilities, people around me regularly engage in activity that supports children to be physically active)

0	1	2	3	4	5	6	7	8	9	10

☐	☐	☐	☐	☐	☐	☐	☐	☐	☐	☐

5. I am motivated to support children and their families in [neighbourhood] to be physically active.

**What is motivation?**

Conscious planning and evaluations (beliefs about what is good and bad about supporting children and their families to be physically active)

(e.g. valuing supporting children’s physical activity, thinking that the potential positive impacts of supporting children to be physically active outweigh any negative impacts)

0	1	2	3	4	5	6	7	8	9	10

☐	☐	☐	☐	☐	☐	☐	☐	☐	☐	☐

6. Supporting children and their families in [neighbourhood] to be physically active is something that I do automatically.

**Automatic motivation** involves doing something (i.e. activities that support children and their families to be physically active) without thinking about it

(e.g. engaging in activities that support children and their families to be active without thinking about it, having a clear plan of tasks to complete)

0	1	2	3	4	5	6	7	8	9	10

☐	☐	☐	☐	☐	☐	☐	☐	☐	☐	☐

7. I am PHYSICALLY able to support children and their families in [neighbourhood] to be physically active.

**What is PHYSICAL capability?**

Having the physical skills required to effectively support children and their families to be physically active

(e.g. sufficient energy, sufficient mobility)

0	1	2	3	4	5	6	7	8	9	10

☐	☐	☐	☐	☐	☐	☐	☐	☐	☐	☐

8. I am PSYCHOLOGICALLY able to support children and their families in [neighbourhood] to be physically active.

**What is PSYCHOLOGICAL capability?**

Knowledge and/or psychological skills to engage in the activities that effectively support children and their families to be physically active

(e.g. knowing how to support children to be active, having sufficient cognitive and interpersonal skills to engage in supporting children to be active).

0	1	2	3	4	5	6	7	8	9	10

☐	☐	☐	☐	☐	☐	☐	☐	☐	☐	☐
