## Additional file 3 for "A whole system approach to increasing children’s physical activity in a multi-ethnic UK city: a process evaluation protocol"

| **JU:MP Bradford Local Delivery Pilot**  **Programme Plan Accelerator Phase 2020 – 2024** | | | | |
| --- | --- | --- | --- | --- |
| **Work stream** | | **Description** | **Themes** | **Lead/Delivery**  **Approach** |
| **OVERARCHING WORKSTREAMS** | | | | |
| **1** | **Strategic Influence** | - Active Bradford (AB) Board and JU:MP Programme Director influence and advocate for the importance of Physical Activity (PA) widely across partnerships. - Sustain the Physical Activity Strategic Development group bringing Living Well (LW) and AB together to collaborate to drive action around PA systems change. - Align the JU:MP Programme to ensure it supports and enhances existing programmes, strategies and plans. - Influence senior leaders, including the Health and Wellbeing board to strengthen PA in policy and practice. - Demonstrate proof of concept of PA whole systems working. | Strategy and policy | JU:MP Programme Director  AB Board |
| **2** | **Workforce**  **Development** | - Develop and deliver training for JU:MP providers to increase knowledge, skills and confidence to increase C&F’s PA using a behaviour change approach. - Deliver training to develop the knowledge, skills and confidence of the wider children’s workforce to increase PA in C&Fs. | Organisations | Core team – CEMS & Programme  Director |
| **3** | **Social Marketing** | - Create a social movement to inspire, educate, and energise children and families (C&F) to be active.   Join Us: Move. Play has four strands:  1. Join the JU:MP movement - getting families to sign up to the Join Us: Move. Play and receive campaign messages and communications.  2. Children and families are active together - promoting local activities and JU:MP website to families.  3. JU:MP Outdoors - Promotes fun and free ideas for outdoor play and the benefits of outdoor play whatever the weather.  4. Reducing sedentary screen time - Raising awareness of the benefits of limiting screen time.   - JU:MP School Fun Days across all 50 schools to encourage families to join the movement. | Families | Commissioned |
| **4** | **Communicating**  **with Partners** | - Facilitate behaviour change in local organisations by building the skills, confidence and motivation to develop effective communication around children's PA. - Development of a joined up “what’s on” city-wide PA search platform. Gaining buy in to open data standards across all partners in Bradford. - Regular communications and updates on JU:MP to key organisations and strategic stakeholders. Agile use of social media to engage key stakeholders. - Development of the AB website and communications to broaden the organisations involved in the promotion of PA messaging. | Organisations  Strategy and policy | JU:MP Core team & Commissioned |
| **5** | **JU:MP Digital App** | - Digital engagement in PA and wider JU:MP Programme, particularly targeting 10 – 13 year olds. Using gamification and behaviour change approaches. - Digital platform which both drives PA engagement while simultaneously capturing data on the reach of the JU:MP programmes and associated health benefits. | Families  Environment | Commissioned |
| **6** | **JU:MP@Home** | - Develop and promote inclusive resources to encourage C&Fs to be active in the home environment. - Develop further and promote the JU:MP@Home website with new activities and ideas shared by local C&Fs. Provide physical resources where needed to address digital exclusion. | Families  Environment | Core team & Commissioned |
| **CROSS-NEIGHBOURHOOD WORKSTREAMS** | | | | |
| **7** | **JU:MP Leads** | - Engage and train local young people aged 16 – 25 years as JU:MP Leads to deliver and facilitate informal multi-sport/PAs. - Training package of sports qualifications, personal development and leadership. - Community Placements within schools, sports clubs, community centres and Youth Services. - JU:MP Leads act as champions through social media and in community. - Support their development as independent social entrepreneurs. | Community | Commissioned |
| **8** | **Sustainable Sport & PA** | - Support local providers to develop PA programmes that take an Asset-Based Community Development (ABCD) approach to creating sustainable changes in PA in C&Fs. - Develop organised sports/active recreation sessions or clubs with a small charge to participants, to create sustainable self-funded PA opportunities. - Train and support local people and mentor JU:MP Leads to deliver sports/PA activity. - Build the capability, confidence and motivation of families to be active independently and/or form social groups to be active together. - To include: community dance programme, girl’s cricket programme and informal multi-sports programmes. | Organisations  Community  Family | Commissioned VCS |
| **9** | **Active Faith Settings** | - Co-produce a toolkit to support local IRS leaders to create healthier and physically active settings. - Workshops with IRS leaders to build capability, opportunity and motivation to create healthy, physically active Madrassa. - Support local IRS and other faith settings to develop individual plans and provide resources (finance and equipment) to implement plans. - Engage IRS and other faith settings in the community JU:MP Programme. | Strategy and policy Organisations | Active Faith Setting Coordinator (IRS Trailblazer partnership) & JU:MP core team |
| **10** | **Creating Active**  **Schools (CAS)** | - Workshops with school leadership teams and CAS leads to enhance capability, opportunity and motivation to create systems change for PA. - Support schools to plan for systems change; focus on whole-school, in-school policy and vision and integrating PA in strategic plans. - Schools to map current resource and PA opportunities and create bespoke school plans to ensure all aspects of CAS Framework are covered. - Provide resources (finance and equipment) and training to schools to meet the needs of individual schools and the collective JU:MP schools. - School leaders’ active involvement in neighbourhood steering groups and delivery of action plans. - Active Travel officers to support school active travel plans and deliver training. | Strategy and policy Organisations | Core team –  CAS Director  Schools |
| **LOCALLY TAILORED WORKSTREAMS** | | | | |
| **11** | **Local**  **Action Plans** | - Establish JU:MP Neighbourhood Action Groups to co-design an action plan built on JU:MP framework through collaborative workshop process. - Neighbourhood action groups identify and prioritise changes to improve local neighbourhoods for PA, e.g. Diversionary PA, community litter picks, etc. - Community Engagement Managers (CEMs) and JU:MP Connectors support neighbourhood steering groups and engage with wider partners to enact their local action plans. Progress is regularly reviewed by the group. | Strategy and policy Community  Organisations  Environment  Families | Core team – CEMS |
| **12** | **JU:MP Connectors** | - JU:MP Connector role in each neighbourhood taking an ABCD approach to increasing children’s PA. - Build the skills, confidence and motivation of less active families to be active together and independently - “Teach a family to fish approach”. - Family engagement in local greenspace, e.g. fun days, led walks, treasure trails. - Support the delivery of the neighbourhood action plan. - Form local groups of C&Fs to co-produce the neighbourhood action plans and greenspace development plans. - The JU:MP Connectors represent C&Fs in the neighbourhood steering groups. | Families  Community  Organisations  Strategy and policy | Commissioned VCS |
| **13** | **Greenspace Development** | - Development of greenspace to improve the local environment for play and PA - At least 5 greenspace developments in Accelerator neighbourhoods. - Community engagement in greenspace developments. - Develop ‘Friends of’ groups for local parks and greenspaces. | Environment  Community | Bradford  Metropolitan  District Council  (BMDC) & CEMs |
| **14** | **Active Playful Parks** | - Outdoor Adventure Play workers provide regular sessions to engage families in nature play areas and establish independent habits of play. - Play Hubs (containers of play equipment within parks) delivered by playworkers, and volunteers in the long-term. - Peel Park Cycling Hub - provision to hire bikes, ‘Dr Bike’ repair and maintenance, learn to ride cycle training, cycling events, and ride leader training. Provide wider support for cycling across JU:MP area. - JU:MP Fun days in greenspace with PA games and trails, with incentives provided for completion of activity challenges. - JU:MP Digital App engages C&F in parks and greenspaces. - Action groups work on safer, cleaner parks, with actions including litter picks, ASB, and agreeing solutions around dogs in parks. | Environment  Families  Community | Commissioned  & BMDC |
| **15** | **Active Travel & Streets** | - Active Travel Consultant to provide expertise to support development of neighbourhood active travel plans and link to wider strategic active travel plans. - Support development of active travel plans for community and faith/IRS organisations. - Playful streets initiatives to animate streets with temporary designs and playful activities such as chalk walks and rain graffiti. | Environment  Organisations | Commissioned |
