## Additional file 1 for "A whole system approach to increasing children’s physical activity in a multi-ethnic UK city: a process evaluation protocol"

| **TITLE** | | | | **Reported in document Y/N/Unclear** | **Page(s) in doc** |
| --- | --- | --- | --- | --- | --- |
| 1 |  | In the title, identify the document as a realist evaluation | | n/a* |  |
| SUMMARY OR ABSTRACT | | | |  |  |
| 2 |  | Journal articles will usually require an abstract, while reports and other forms of publication will usually benefit from a short summary. The abstract or summary should include brief details on: the policy, programme or initiative under evaluation; programme setting; purpose of the evaluation; evaluation question(s) and/or objective(s); evaluation strategy; data collection, documentation and analysis methods; key findings and conclusions  Where journals require it and the nature of the study is appropriate, brief details of respondents to the evaluation and recruitment and sampling processes may also be included  Sufficient detail should be provided to identify that a realist approach was used and that realist programme theory was developed and/or refined | | Y | 2 |
| INTRODUCTION | | | |  |  |
| 3 | Rationale for evaluation | | Explain the purpose of the evaluation and the implications for its focus and design | Y | 4-5 |
| 4 | Programme theory | | Describe the initial programme theory (or theories) that underpin the programme, policy or initiative | Y | 3, 5-6 |
| 5 | Evaluation questions, objectives and focus | | State the evaluation question(s) and specify the objectives for the evaluation. Describe whether and how the programme theory was used to define the scope and focus of the evaluation | Y (methods) | 8, 10 |
| 6 | Ethical approval | | State whether the realist evaluation required and has gained ethical approval from the relevant authorities, providing details as appropriate. If ethical approval was deemed unnecessary, explain why | Yes (methods) | 8, 10 |
| METHODS | | | |  |  |
| 7 | Rationale for using realist evaluation | | Explain why a realist evaluation approach was chosen and (if relevant) adapted | Y | 7-8 |
| 8 | Environment surrounding the evaluation | | Describe the environment in which the evaluation took place | Y | 4 |
| 9 | Describe the programme policy, initiative or product evaluated | | Provide relevant details on the programme, policy or initiative evaluated | Y | 5-6 |
| 10 | Describe and justify the evaluation design | | A description and justification of the evaluation design (i.e. the account of what was planned, done and why) should be included, at least in summary form or as an appendix, in the document which presents the main findings. If this is not done, the omission should be justified and a reference or link to the evaluation design given. It may also be useful to publish or make freely available (e.g. online on a website) any original evaluation design document or protocol, where they exist | Y | 8-11 |
| 11 | Data collection methods | | Describe and justify the data collection methods – which ones were used, why and how they fed into developing, supporting, refuting or refining programme theory  Provide details of the steps taken to enhance the trustworthiness of data collection and documentation | Y | 12-13 |
| 12 | Recruitment process and sampling strategy | | Describe how respondents to the evaluation were recruited or engaged and how the sample contributed to the development, support, refutation or refinement of programme theory | Y | 11-12 |
| 13 | Data analysis | | Describe in detail how data were analysed. This section should include information on the constructs that were identified, the process of analysis, how the programme theory was further developed, supported, refuted and refined, and (where relevant) how analysis changed as the evaluation unfolded | Y  (plan) | 18-19 |
| RESULTS | | | |  |  |
| 14 | Details of participants | | Report (if applicable) who took part in the evaluation, the details of the data they provided and how the data was used to develop, support, refute or refine programme theory | n/a* |  |
| 15 | Main findings | | Present the key findings, linking them to contexts, mechanisms and outcome configurations. Show how they were used to further develop, test or refine the programme theory | n/a* |  |
| DISCUSSION | | | |  |  |
| 16 | Summary of findings | | Summarise the main findings with attention to the evaluation questions, purpose of the evaluation, programme theory and intended audience | n/a* |  |
| 17 | Strengths, limitations and future directions | | Discuss both the strengths of the evaluation and its limitations. These should include (but need not be limited to): (1) consideration of all the steps in the evaluation processes; and (2) comment on the adequacy, trustworthiness and value of the explanatory insights which emerged  In many evaluations, there will be an expectation to provide guidance on future directions for the programme, policy or initiative, its implementation and/or design. The particular implications arising from the realist nature of the findings should be reflected in these discussions | 1Y  2n/a* |  |
| 18 | Comparison with existing literature | | Where appropriate, compare and contrast the evaluation’s findings with the existing literature on similar programmes, policies or initiatives | n/a* | 20-21 |
| 19 | Conclusion and recommendations | | List the main conclusions that are justified by the analyses of the data. If appropriate, offer recommendations consistent with a realist approach | n/a* |  |
| 20 | Funding and conflict of interest | | State the funding source (if any) for the evaluation, the role played by the funder (if any) and any conflicts of interests of the evaluators | Y | 22 |

*This study is not a realist evaluation but rather is informed by realist principles. Therefore, it was not applicable to include the terminology ‘realist evaluation’ in the title. Additionally, this manuscript reports on a process evaluation protocol, meaning there are no results presented, and so it was not applicable to include many of the items in the results and discussions sections of the reporting template. There is no reporting tool available for the reporting of process evaluations or process evaluation protocols and the RAMSES reporting template was identified as the most applicable available template after reviewing other reporting templates that we considered could be applicable e.g. SPIRIT (reporting protocols of clinical trials) and STaRI (reporting implementation studies).
