## Additional file 7 for "A whole system approach to increasing children’s physical activity in a multi-ethnic UK city: a process evaluation protocol"

**A process evaluation of the strategic and neighbourhood-level design, delivery, and evaluation of the JU:MP programme**

Focused on understanding the views and actions of the strategic team and neighbourhood action groups in relation to: (Part A) Design and delivery of the overarching and neighbourhood JU:MP programme, (Part B) Design and delivery of specific JU:MP programmes / projects / commissions (Part C) JU:MP research and evaluation.

Objectives:

(1) To document the strategic-level design, delivery and evaluation processes of the JUMP programme
(2) To examine the *feasibility* and *acceptability* of the strategic level design, delivery and evaluation of the JU:MP programme by understanding the barriers and facilitators and contextual factors influencing design, delivery and evaluation
(3) To understand the *impact* of the JUMP programme including developing an understanding of what works, for whom, and in what context

**Part A: The overarching JU:MP programme**

This section relates to the overarching JU:MP programme, including, for example, the overarching approach being taken / aims / theory, themes, commissioning processes, and the neighbourhood approach to delivery of the programme, intervention development processes, the research-practice partnership approach. Data that relates to discrete JU:MP work streams should be coded in Part B and data that relates to evaluation work packages should be coded in Part C.

| **Subtheme name** | **Subtheme description** | **Links to theory** |
| --- | --- | --- |
| 1.1 Overarching programme design, characteristics and acceptability | | |
| 1.1.1 Knowledge about the overarching JU:MP programme | Knowledge about the overarching programme e.g. whole system approach, aims including sustainability, theory, neighbourhood approach to delivery, themes, commissioning process, research-led programme. resource/cost, level of complexity / adaptability / replicability. This refers to anything the stakeholder perceives the project to be ‘about’ - still code here if inaccurate perception. | TDF - knowledge  CFIR - knowledge about the intervention; cost / resource; adaptability / complexity, design quality and packaging |
| 1.1.2. Source of the JU:MP programme and strength of the underpinning evidence | Perceptions of interviewees about how and by whom the overarching programme has been developed / is being developed, including for example: development of ToC and implementation plan, evidence-based, theoretical basis, co-production of the approach / plan (source); Perceptions about the quality and validity of the evidence supporting the overarching programme; strength of the rationale / problem and proposed mechanisms of action of the programme (strength of evidence). **Exclude iterative development based on** **process evaluation and learning (code to 1.4).** | CFIR - intervention source, strength of the evidence |
| 1.1.3 Feelings about the overarching JU:MP programme - positive | Positive feelings about the programme including what the stakeholder likes about the JU:MP programme (incl compared to other solutions), perceptions about what ‘works well’ and why - about the programme design not implementation (e.g. complexity, adaptability, neighbourhood approach). **Exclude: impact - i.e. any change (code to 1.3), reflection re what works well that leads to adapted practice (code to 1.4).** | Affective attitude (Sekhon)  CFIR - beliefs about the intervention, design quality and packaging, adaptability, complexity |
| 1.1.4. Feelings about the overarching JUMP programme - negative | Including what the stakeholder doesn’t like about the JU:MP programme (including compared to alternative solutions), why, and what they would change about it or do differently in future - **about the programme design not implementation. Exclude: impact - i.e. any change (code to 1.3), reflection on implementation that leads to adapted practice (code to 1.4)** | Affective attitude (Sekhon)  CFIR - beliefs about the intervention, design, design quality and packaging, adaptability, complexity |
| 1.1.5. Connection / synergy / tension between different elements of the programme | Detail re where different components of the programme (e.g. different interventions) experience connection, synergy or tension, including the extent to which and how this is explored as part of the overarching JU:MP programme **(at a design level - code implementation factors to 1.2.3.5)** | Systems |
| 1.2 Implementation of the overarching programme | | |
| 1.2.1 Implementation activity (core team) - programme development | Planning, engagement and execution related to the overarching programme development / delivery, eg. commissioning, developing neighbourhoods, developing the JU:MP programme model, theory of change. **Exclude content that relates to a specific workstream.** | CFIR - process  NPT |
| 1.2.2 Implementation activity (core team) - programme evaluation | Planning, engagement and execution related to the overarching programme evaluation, e.g. outcomes framework. **Exclude content that relates to a specific evaluation work package.** | CFIR - process  NPT |
| 1.2.3 Factors influencing whether core team can deliver role generally (not specific to a work stream or research package) / implementation of the overarching programme: inner setting | | |
| 1.2.3.1 Capability of core team (members) to deliver the JU:MP programme and evaluation | The individual's knowledge about topics relevant to their role, and previous experiential learning (prior to JU:MP). Actual (skills, decision making) and perceived capability of the individual to complete their role in delivering workstream. Include relevant previous experience and thoughts about training required. Note - this can be implicit or explicit. **Exclude: statements about what they think JU:MP is, as this is covered in 1.1.1; Exclude content that relates to a specific evaluation work package.** | TDF - knowledge, skills, beliefs in capability, memory / attention  Sekhon - self-efficacy |
| 1.2.3.2 Motivation of core team (members) to deliver the JU:MP programme and evaluation | Personal identity / values - the importance of delivering their JU:MP role; extent to which work stream aligns with personal values; the extent to which values must be given up to engage in a role. How JU:MP makes them feel; how emotion /mental health affects their work. **Exclude content that relates to a specific evaluation work package.** | TDF - intention, identity, emotion  Sekhon - opportunity costs, ethicality  CRIF - compatibility |
| 1.2.3.3 Core JU:MP team context, including deliver organisation factors | Factors related to the core JU:MP team that make it easier / harder to deliver the overall programme(incl evaluation). Including: social architecture, age, maturity, size etc. of the JU:MP team, ‘culture’ of the team (including whether they feel valued as part of the team, safe to try new methods etc.). The environmental context of the inner / core JU:MP team including networks and communication processes, the physical environment (office space etc.); access to resources. **Exclude content that relates to a specific evaluation work package.** | TDF - social env; env context and resource  CFIR - inner setting; structural characteristics; impl climate, relative priority, networks + comms; resources |
| 1.2.3.4 Programme leadership, strategic direction | Influence of programme leadership and strategic direction on the implementation of the overarching JU:MP programme. Including the extent to which leaders motivate the team, AB board, JU:MP exec board, governance processes etc. |  |
| 1.2.3.5 Influence of funding body | Influence of the funding organisation (i.e. Sport England) on implementation of the programme, e.g. guidance provided by funder, community perceptions of funder. | CFIR - inner setting |
| 1.2.4 Factors influencing whether core team can deliver role generally (not specific to a work stream or research package) / implementation of the overarching programme: outer setting | | |
| 1.2.4.1 Factors related to commissioned organisations / delivery stakeholders | Factors related to commissioned orgs / delivery stakeholders, that influence implementation of the overarching programme, including feelings about and engagement with the JU:MP programme (and evaluation), barriers and facilitators to supporting delivery - **where it doesn't relate to a specific work stream or evaluation component.** | COM-B / TDF  CFIR - patient needs and resources |
| 1.2.4.2 Factors related to local children and families | Factors related to local children and families that influence implementation of the overarching programme, including children / family feelings about and engagement with the JU:MP programme (and evaluation), barriers and facilitators to being physical activity and engaging in evaluation - **where it doesn't relate to a specific work stream or evaluation component.** | COM-B / TDF  CFIR - patient needs and resources |
| 1.2.4.3 Factors related to local organisations and wider stakeholders | Factors related to local organisations and wider stakeholders that influence implementation of the overarching programme, Including feelings about and engagement with the JU:MP programme (and evaluation), barriers and facilitators to supporting delivery / changing behaviour - **where it doesn't relate to a specific work stream or evaluation component.** | COM-B / TDF  CFIR - patient needs and resources |
| 1.2.4.4 Bradford area context (including local policy / strategy) | Factors related to the nature / characteristics of the LDP area and wider Bradford area, that influence implementation of the overarching programme and, including: Structural characteristics, culture, environmental context and resources such as greenspace / local assets, demographics of residents, history, policies and local politics etc. The degree to which JU:MP is networked with other Bradford programmes. | CFIR - structure, networks, culture, impl climate, leadership engagement, available resources  CFIR - outer setting; cosmopolitanism |
| 1.2.4.5 Wider contextual factors | Wider (regional / (inter)national) social, cultural, political, societal, governmental, environmental factors that influence delivery of the work stream e.g. partnerships with wider programmes e.g. other LDPs, COVID-19, political purdah | CFIR - outer setting; external policies and incentives, peer pressure; cosmopolitanism |
| 1.2.5 Challenges, considerations, reflection and learning in key areas of overarching programme development, delivery and evaluation | | |
| 1.2.5.1 Hyperlocal neighbourhood approach, bottom up / top down | Challenges, considerations, reflection and learning specific to the delivery approach - hyperlocal approach vs ‘top down’ interventions, community engagement / co-production / ABCD; doing ‘to’ vs doing ‘with’; managing working in different neighbourhoods, challenges of selecting neighbourhoods etc. | CFIR - intervention characteristics, process (reflection) |
| 1.2.5.2 Preparing accelerator bid and developing the JU:MP model… | Challenges, considerations, reflection and learning related to the process of preparing the accelerator bid and developing the JU:MP model **(Exclude references to hyperlocal approach/selecting neighbourhoods etc., code to 1.2.3.1)** | CFIR - intervention characteristics,  Process (reflection) |
| 1.2.5.3 Staffing structure, role clarity, (internal) communication | Challenges, considerations, reflection and learning related to the JU:MP core team staffing structure, role clarity, and (internal) communications. | CFIR - intervention characteristics,  Process (reflection), inner setting |
| 1.2.5.4 Research-practice partnership and evaluation | Challenges, considerations, reflection and learning related to the research-practice partnership approach to delivering JU:MP, and the overarching evaluation. | CFIR - intervention characteristics,  Process (reflection) |
| 1.2.5.5 Whole system programme delivery | Challenges, considerations, reflections and learning related to delivering a whole-system programme with multiple interacting components, including, synergies and tensions related to delivery, timings of different elements, programme management considerations etc. | CFIR - intervention characteristics,  Process (reflection) |
| 1.2.5.6 Commissioning and allocation of resource | Challenges, considerations, reflections and learning related to commissioning and allocation of resources as part of the overarching JU:MP programme. | CFIR - costs, intervention characteristics,  Process (reflection) |
| 1.3 Impact, mechanisms of impact and wider (systems) change | | |
| Note that this theme relates to JU:MP overall, or when talking about the impact of work streams in combination, or the impact of JU:MP within neighbourhoods. Workstream specific - code within workstream theme. Research and evaluation specific - code there. | | |
| 1.3.1 Beliefs about consequences | Expected / anticipated (not actual) consequences of JU:MP; the extent to which JU:MP is perceived as likely to achieve its purpose; positive / negative. | TDF - beliefs about conseq  Sekhon - perceived effectiveness |
| 1.3.2. Influence of JU:MP on children and young people | Interviewees perception of the impact (or lack thereof) of JU:MP on C&YP, in terms of their behaviour (physical activity) as well as on factors that directly influence C&YP behaviour, e.g. their capability, opportunity, or motivation to engage in PA, differing impact on different groups and reasons for this. | Theory of change  COM-B / TDF  Systems |
| 1.3.3. Influence of JU:MP on families | Interviewees perception of the impact (or lack thereof) of JU:MP on families (e.g. parents, guardians, siblings, other family members), in terms of their behaviour (physical activity, supporting / encouraging children to be active) as well as on factors that directly influence their behaviour, e.g. their capability, opportunity, or motivation to engage in PA or support children to engage in PA, differing impact on different groups and reasons for this. | Theory of Change  COM-B / TDF  Systems |
| 1.3.4. Influence of JU:MP on organisations | Interviewee’s perception of the impact (or lack thereof) of JU:MP on organisations, in terms of the behaviour of organisational stakeholders (encouraging children to be active) and incorporating PA in policies / practices, as well as on factors that directly influence organisational behaviour, e.g. their capability, opportunity, or motivation to support children to engage in PA; differing impact on different groups and reasons for this. | Theory of Change  COM-B / TDF  Systems |
| 1.3.5. Influence of JU:MP on stakeholders involved in delivery | Interviewee’s perception of the impact (or lack thereof) of JU:MP on strategic / neighbourhood deliverers, e.g. those who are leading on programmes at whole programme or neighbourhood level such as CEMs, active travel leads, researchers, exec board, policy and strategy leads, school stakeholders, JU:MP leads, connectors, differing impact on different groups and reasons for this. | COM-B / TDF  Theory of Change  Systems |
| 1.3.6. Perceived influence of wider (non-JUMP) factors or activities on the impact of JU:MP | Interviewee’s perception of non-JUMP activities (i.e. moderators) on the success of the workstream including moderators that may interact with JU:MP to constrain and / or facilitate success, or that independently contribute to changes in relevant outcomes. | Theory of change  Systems |
| 1.3.7. Wider outcomes of work stream - safer more active communities | Wider outcomes of JU:MP linked to communities / the neighbourhood system, including for example, more ‘joined up’ / connected communities, environmental outcomes, safer neighbourhoods. | Theory of Change  Systems |
| 1.3.8. Wider outcomes of work stream - individual C&YP | Wider outcomes specific to the individual end users (C&YP) including better skills, better physical health, and better mental health. | Theory of change  Systems |
| 1.3.9. Unintended negative consequences | Actual consequences of JU:MP that were unintended and negative (not part of theory of change / linked to desired outcomes) e.g. negative perceptions about JU:MP which lead to disengagement, exacerbation of health inequalities through interventions ‘working’ better for some groups than others. | Systems |
| 1.3.10 Impact of JU:MP research and evaluation activity | The influence of the broad research / evaluation activity, including, for example, on the attitudes or behaviours of key stakeholders, including the JU:MP team, steering groups, organisation, children and families. **Exclude where impact relates to specific activity e.g impact of specific evaluation workpackage, impact on specific work stream** | Systems  Theory of Change  COM-B / TDF |
| 1.4 General reflection and learning | | |
| 1.4.1 Reflection and learning (positive) | Including interviewee perceptions related to ‘what worked’ and why with regards to the overarching programme delivery, and impact on iterative programme development. **Where it does not fit within any of the sub-themes within 1.2.3.** | Systems  CFIR - process |
| 1.4.2 Reflection and learning (negative) | Including interviewee perceptions related to ‘what didn't work’ in implementing the overarching programme and why, what they would do differently in future, and impact on iterative programme development. **Where it does not fit within any of the sub themes within 1.2.3.** | Systems  CFIR - process |
| 1.4.3 Perceptions about reflection and learning | Perceptions about and instances of opportunities for reflection and learning in relation to the overarching programme (e.g. the way we work), utility of reflection, perception re whether reflection leads to change in practice | CFIR - process  NPT  Systems |

**Part B: JU:MP work streams**

This section relates to the discrete work streams that JU:MP comprises - there is a separate theme for each work stream. Data that is specific to a work stream should be coded into the respective theme.

Theme 2: Strategic Influencing; Theme 3: Communicating with partners; Theme 4: Social Marketing; Theme 5: Workforce development; Theme 6: JUMP@Home; Theme 7: The JU:MP digital App; Theme 8: Creating Active Schools; Theme 9: Active Faith Settings; Theme 10: JU:MP Leads; Theme 11: Sustainable Sport and Physical Activity; Theme 12: Neighbourhood Action Plans; Theme 13: Greenspace development; Theme 14: Active Playful Parks; Theme 15: Active Travel and Streets; Theme 16: JU:MP Connectors.

**The neighbourhood action group plans work stream theme should include the neighbourhood action group approach.**

*For each theme in part B, the subthemes to be as follows:*

| X.1 Workstream design, characteristics and acceptability | | |
| --- | --- | --- |
| x.1.1. Knowledge about workstream | Knowledge about work stream - aims, scope, ethos, complexity, adaptability, replicability etc. This refers to anything the stakeholder *perceives* the project to be ‘about’ - still code here if inaccurate according to investment plan / other documents. | TDF - knowledge  CFIR - knowledge about the intervention, adaptability, complexity |
| X.1.2. Source of workstream and strength of underpinning evidence | Perceptions about how and by whom the workstream has been developed / is being developed, including for example: development of ToC and implementation plan, evidence-based, primary research, theoretical basis, co-production of the approach / plan, planning activity; Perceptions about the quality and validity of the evidence supporting the project; strength of the rationale / problem and proposed mechanisms of action of the project (strength of evidence). **Exclude: iterative development based on reflection / learning from implementation and evaluation (code to x.4.8 or x.4.9).** | CFIR - intervention source, evidence strength and quality |
| x.1.3. Feelings about workstream | Including what they like and do not like about the work stream, what ‘works well’ and why i.e. the ’activities’ that lead to change. **Exclude: impact (code to x.3), reflection that leads to adapted practice (code to x.4)** | Affective attitude (Sekhon)  CFIR - beliefs about the intervention |
| x.1.4. Compatibility with the wider JU:MP approach | Detail re any connection, synergy or tension between the work stream and the values / principles of the overarching JU:MP programme or other work streams, in terms of design. **Exclude: tensions / compatibility related to implementation (code to x.2.3.6)** | Systems |
| X.2 Workstream implementation | | |
| x.2.1. Implementation activity (workstream leads / core team) - engagement; cognitive participation; execution | Workstream strategic implementation activity - Engagement of leads, including how strategic leads are trained / engaged to deliver the work stream; implementation planning; carrying out / accomplishing implementation according to plan (what happened, when, how, by whom etc) | CFIR - implementation  NPT |
| x.2.2 Implementation activity (org /stakeholders - not strategically leading but working across LDP rather than within neighbourhood) | Workstream implementation activity completed by organisational or other stakeholders across (not within) neighbourhoods - Engagement of stakeholders, including how stakeholders are trained / engaged to deliver the work stream; implementation planning; carrying out / accomplishing implementation according to plan (what happened, when, how, by whom etc) | CFIR - implementation  NPT |
| X.2.3 Workstream implementation within JU:MP neighbourhoods | | |
| x.2.3.1 PN A (AllerGrange) | Workstream neighbourhood implementation activity - Engagement of neighbourhood stakeholders, including community engagement; planning; carrying out / accomplishing implementation according to plan (what happened, when, how, by whom etc) | CFIR - implementation  NPT |
| X.2.3.2 - PN B (Scotchman Rd) | Workstream neighbourhood implementation activity - Engagement of neighbourhood stakeholders, including community engagement; planning; carrying out / accomplishing implementation according to plan (what happened, when, how, by whom etc) | CFIR - implementation  NPT |
| x.2.3.3 - PN C  (Peel Park) | Workstream neighbourhood implementation activity - Engagement of neighbourhood stakeholders, including community engagement; planning; carrying out / accomplishing implementation according to plan (what happened, when, how, by whom etc) | CFIR - implementation  NPT |
| X.2.3.4 Delivery neighbourhood context - DDN A | Workstream neighbourhood implementation activity - Engagement of neighbourhood stakeholders, including community engagement; planning; carrying out / accomplishing implementation according to plan (what happened, when, how, by whom etc) | CFIR - implementation  NPT |
| X.2.3.5 Delivery neighbourhood context - DDN B | Workstream neighbourhood implementation activity - Engagement of neighbourhood stakeholders, including community engagement; planning; carrying out / accomplishing implementation according to plan (what happened, when, how, by whom etc) | CFIR - implementation  NPT |
| X.2.3.6 Delivery neighbourhood context - DDN C | Workstream neighbourhood implementation activity - Engagement of neighbourhood stakeholders, including community engagement; planning; carrying out / accomplishing implementation according to plan (what happened, when, how, by whom etc) | CFIR - implementation  NPT |
| X.2.3.7 Delivery neighbourhood context - Commissioned neighbourhood A | Workstream neighbourhood implementation activity - Engagement of neighbourhood stakeholders, including community engagement; planning; carrying out / accomplishing implementation according to plan (what happened, when, how, by whom etc) | CFIR - implementation  NPT |
| X.2.3.8 Delivery neighbourhood context - Commissioned neighbourhood B | Workstream neighbourhood implementation activity - Engagement of neighbourhood stakeholders, including community engagement; planning; carrying out / accomplishing implementation according to plan (what happened, when, how, by whom etc) | CFIR - implementation  NPT |
| X.2.4 ‘Inner setting’ factors influencing workstream implementation | | |
| x.2.4.1. ‘Deliverer’ (individual) (belief in) capability to deliver workstream | The individual's knowledge about relevant topics, including *perceptions* about their level of understanding of the workstream, and previous experiential learning (prior to JU:MP). Actual (skills, decision making) and perceived capability of the individual to complete their role in delivering workstream. Include relevant previous experience and thoughts about training required. Note - this can be implicit or explicit. **Exclude: statements about what they think the workstream is, code to x.1.1.** | TDF - knowledge, skills, beliefs in capability, memory / attention  Sekhon - self-efficacy |
| X.2.4.2 ‘Deliverer’ (individual) motivation to deliver workstream | Personal identity / values - the importance of delivering their JU:MP role (where related to specific work stream); extent to which work stream aligns with personal values; the extent to which values must be given up to engage in a role. How JU:MP / the person's role in delivering the work stream makes them feel; how emotion /mental health affects their work. The extent to which JU:MP aligns with professional role outside of JU:MP. | TDF - intention, identity, emotion  Sekhon - opportunity costs, ethicality  CRIF - compatibility |
| x.2.4.3. Commissioned organisations | Factors related to the nature / characteristics of organisations that are commissioned to deliver the JU:MP work stream, that make it easier / harder to deliver the workstream, including:  Structural characteristics, culture (incl in relation to whether values align with those of JU:MP, and extent to which JUMP work is prioritised), environmental context and resources, policies and processes, communication and way of working with the JU:MP team, including how communication from within core JUMP team affects commissioned orgs/other strategic-level stakeholders. | TDF - Env context and resources  CFIR - inner setting; impl climate, compatibility, structural characteristics, culture, relative priority, learning climate, comms / networks, resources |
| x.2.4.4. Characteristics of the work stream itself | Characteristics of the workstream itself that make it easier / harder to implement. | CRIF - intervention characteristics |
| X.2.4.5 Elements of the wider JU:MP programme | Characteristics of the wider JU:MP programme, including other work streams and the research/evaluation, that makes the workstream easier / harder to implement. | CFIR - inner setting |
| x.2.4.6. Core JU:MP team context (physical, social, cultural, organisational) | Factors related to the core JU:MP team that make it easier / harder to deliver the workstream. Including:  Social architecture, age, maturity, size etc. of the JU:MP team, ‘culture’ of the team (including whether feel valued as part of the team, safe to try new methods etc.).  The environmental context of the inner / core JU:MP team including networks and communication processes, the physical environment (office space etc.) access to resources | TDF - social env; env context and resource  CFIR - inner setting; structural characteristics; impl climate, relative priority, networks and comms; resources |
| x.2.4.7. Influence of JU:MP funding body | Influence of the funding organisation (i.e. Sport England) on implementation of the work stream, e.g. guidance provided by funder, perceptions of funder. **Exclude: resource allocated to work stream (code to x.2.3.5)** | CFIR - inner setting |
| x.2.4.8. Programme leadership | Programme leadership factors influencing delivery of the work stream, including: commitment, involvement, and accountability of leaders and managers, including the extent to which leaders inspire / motivate the team; including team leads, AB board, JU:MP exec board. Incentives for deliverers such as goal-sharing awards, performance reviews / goal setting and feedback processes, promotions, and less tangible incentives such as increased stature or respect | CFIR - inner setting; leadership engagement; organisational incentives and rewards; goals and feedback |
| X.2.5 ‘Outer setting’ factors influencing workstream implementation | | |
| x.2.5.1. Factors related to local children and families | Including children/family need for the work stream / feelings about work stream, COM of children families to engage with work stream, barriers / facilitators to engaging children & families in this work stream. | COM-B / TDF  CFIR - patient needs and resources |
| x.2.5.2. Factors related to local organisations (e.g. schools, IRS, VCS) | Including perceived need for work stream / thoughts about work stream (from perspective of organisation), COM of organisation for supporting/ receiving work stream, barriers / facilitators to engaging with organisational related to this work stream. | COM-B / TDF  CFIR - patient needs and resources |
| x.2.5.3. Factors related to other stakeholders | Including wider stakeholder perceived need for work stream / thoughts about work stream, COM of other stakeholders for supporting JU:MP, barriers / facilitators to engaging with other stakeholders related to this work stream. Stakeholders may include residents, local leaders, councillors, policy-makers. | COM-B/TDF  CFIR - patient needs and resources |
| x.2.5.4. Bradford area context (including local policy/strategy) | Factors related to the nature / characteristics of the LDP area and wider Bradford area, that influence implementation of the work stream, including:  Structural characteristics, culture, environmental context and resources such as greenspace / local assets, demographics of residents, history, policies and local politics etc. The degree to which JU:MP is networked with other Bradford programmes or organisations | CFIR - inner context; structure, networks, culture, impl climate, leadership engagement, available resources  CFIR - outer setting; cosmopolitanism |
| x.2.5.5. Wider contextual factors | Wider (regional / (inter)national) social, cultural, political, societal, governmental, environmental factors that influence delivery of the work stream e.g. partnerships with wider programmes e.g. other LDPs, COVID-19, political purdah | CFIR - outer setting; external policies and incentives, peer pressure; cosmopolitanism |
| X.2.5.6. Delivery neighbourhood context - factors influencing work stream implementation | | |
| x.2.5.6.1 PN A (AllerGrange) | Factors related to the nature / characteristics of PN A, that influence delivery of the work stream, including:  Structural characteristics (size), culture, environmental context and resources such as greenspace / local assets, demographics of residents, history, how ‘joined up’ local stakeholders are, community engagement etc. | CFIR - inner context; structure, networks, culture, impl climate, resources  CFIR - outer setting; needs |
| x.2.5.6.2 - PN B (Scotchman Rd) | Factors related to the nature / characteristics of PN B, that influence delivery of the work stream, including:  Structural characteristics (size), culture, environmental context and resources such as greenspace / local assets, demographics of residents, history, how ‘joined up’ local stakeholders are, community engagement etc. | CFIR - inner context; structure, networks, culture, impl climate, available resources  CFIR - outer setting; needs |
| x.2.5.6.3 - PN C  (Peel Park) | Factors related to the nature / characteristics of PN C, that influence delivery of the work stream, including:  Structural characteristics (size), culture, environmental context and resources such as greenspace / local assets, demographics of residents, history, how ‘joined up’ local stakeholders are etc. | CFIR - inner context; structure, networks, culture, impl climate, available resources  CFIR - outer setting; needs |
| X.2.5.6.4 Delivery neighbourhood context - DDN A | Factors related to the nature / characteristics of DDN A, that influence delivery of the work stream, including:  Structural characteristics (size), culture, environmental context and resources such as greenspace / local assets, demographics of residents, history, how ‘joined up’ local stakeholders are etc. | CFIR - inner context; structure, networks, culture, impl climate, available resources  CFIR - outer setting; needs |
| X.2.5.6.5 Delivery neighbourhood context - DDN B | Factors related to the nature / characteristics of DDN B, that influence delivery of the work stream, including:  Structural characteristics (size), culture, environmental context and resources such as greenspace / local assets, demographics of residents, history, how ‘joined up’ local stakeholders are etc. | CFIR - inner context; structure, networks, culture, impl climate, available resources CFIR - outer setting; need |
| X.2.5.6.6 Delivery neighbourhood context - DDN C | Factors related to the nature / characteristics of DDN C, that influence delivery of the work stream, including:  Structural characteristics (size), culture, environmental context and resources such as greenspace / local assets, demographics of residents, history, how ‘joined up’ local stakeholders are etc. . | CFIR - inner context; structure, networks, culture, impl climate, available resources  CFIR - outer setting; need |
| X.2.5.6.7 Delivery neighbourhood context - Commissioned neighbourhood A | Factors related to the nature / characteristics of commissioned neighbourhood A, that influence delivery of the work stream, including:  Structural characteristics (size), culture, environmental context and resources such as greenspace / local assets, demographics of residents, history, how ‘joined up’ local stakeholders are etc. | CFIR - inner context; structure, networks, culture, impl climate, available resources  CFIR - outer setting; need |
| X.2.5.6.8 Delivery neighbourhood context - Commissioned neighbourhood B | Factors related to the nature / characteristics of commissioned neighbourhood B, that influence delivery of the work stream, including:  Structural characteristics (size), culture, environmental context and resources such as greenspace / local assets, demographics of residents, history, how ‘joined up’ local stakeholders are etc. | CFIR - inner context; structure, networks, culture, impl climate, available resources  CFIR - outer setting; need |
| X.3. Workstream impact, mechanisms of change, and wider (system) change | | |
| x.3.1. Beliefs about consequences | Expected / anticipated (not actual) impact of the work stream; the extent to which the work stream is perceived as likely to achieve its purpose; positive / negative. | TDF - beliefs about consequences  Sekhon - perceived effectiveness |
| x.3.2. Influence of work stream on children and young people | Perception of the impact (or lack thereof) of the work stream on C&YP’s physical activity, as well as on factors that directly influence C&YP behaviour, e.g. their capability, opportunity, or motivation to engage in PA, differing impact on different groups and reasons for this. | Theory of change  COM-B / TDF  Systems |
| x.3.3. Influence of work stream on families | Perception of the impact (or lack thereof) of the work stream on families (e.g. guardians, siblings, other family members),behaviour (physical activity, supporting / encouraging children to be active) as well as on factors that directly influence their behaviour, e.g. their capability, opportunity, or motivation; differing impact on different groups and reasons for this. | Theory of Change  COM-B / TDF  Systems |
| x.3.4. Influence of work stream on organisations | Interviewees perception of the impact (or lack thereof) of the work stream on organisation behaviour (e.g. encouraging children to be active, incorporating PA in policies / practices), as well as on factors that directly influence organisational behaviour, e.g. their capability, opportunity, or motivation; differing impact on different groups and reasons for this. | Theory of Change  COM-B / TDF  Systems |
| x.3.5. Influence of work stream on stakeholders involved in delivery | Interviewees perception of the impact (or lack thereof) of the work stream on strategic / neighbourhood deliverers, e.g. those who are leading on programmes at whole programme or neighbourhood level such as CEMs, active travel leads, researchers, exec board, policy and strategy leads, school stakeholders, JU:MP leads, connectors, differing impact on different groups and reasons for this. | COM-B / TDF  Theory of Change  Systems |
| x.3.6. Perceived influence of wider (non-JUMP) factors on the impact of the work stream | Interviewees perception of non-JUMP activities (i.e. moderators) on the success of the work stream including moderators that may interact with JU:MP to constrain and / or facilitate success, or that independently contribute to changes in relevant outcomes. | Theory of change  Systems |
| x.3.7. Wider impact of work stream - safer more active communities | Wider impact of the work stream linked to communities / the neighbourhood system, including for example, more ‘joined up’ / connected communities, environmental outcomes, safer neighbourhoods. | Theory of Change  Systems |
| x.3.8. Wider impact of work stream - C&YP | Wider impact of work stream specific to C&YP including better skills, better physical health, better mental health. | Theory of change  Systems |
| x.3.9. Unintended negative consequences | Consequences of the work stream that were unintended and negative e.g. perceptions about JU:MP which lead to disengagement, exacerbation of health inequalities | Systems |
| X.4 Research, evaluation, reflection and learning | | |
| X.4.1 Knowledge / attitudes related to workstream research / evaluation activity | Knowledge about JU:MP workstream research / evaluation; this refers to anything the stakeholder *perceives* the project to be ‘about’ - still code here if inaccurate. Attitudes about work stream-specific research / evaluation activity including what they like and do not like about the research activity, what ‘works well’ and why. **Exclude: impact (code to x.4.6/8)** | TDF - knowledge  CFIR - knowledge and beliefs about the intervention;  Affective attitude (Sekhon) |
| X.4.2 Implementation of workstream research / evaluation activity | Workstream research /evaluation implementation activity - engagement of leads, including how strategic leads are trained / engaged to deliver the work stream evaluation; planning; carrying out / accomplishing implementation of research / evaluation according to plan (what happened, when, how, by whom etc) | CFIR - implementation  NP |
| X.4.3 Factors influencing workstream research implementation - inner setting | ‘Inner setting’ factors influencing workstream research / evaluation implementation, including deliverer capability motivation and other personal attributes, workstream or research characteristics, characteristics of the JU:MP programme, team context (commitment of wider JU:MP team to supporting research), leadership, funding body. | CFIR - inner setting  COM-B / TDF |
| X.4.4 Factors influencing workstream research implementation - outer setting | ‘Outer setting’ factors influencing workstream research / evaluation implementation, including local children and families (including the perceived feasibility and acceptability of the research), organisations, neighbourhood context, Bradford area context, wider contextual factors | CFIR - outer setting  COM-B / TDF |
| X.4.5 Connectedness of workstream research and implementation | How connected the work stream research and implementation are, including reference to processes, communication, collaboration etc. | CFIR - inner setting  COM-B / TDF |
| X.4.6 Impact of workstream research / evaluation activity | The influence of the research / evaluation activity, including, for example, on the attitudes or behaviours of key stakeholders, including the JU:MP team, steering groups, organisation, children and families. | Systems  Theory of Change  COM-B / TDF |
| X.4.7 Reflection and learning (positive) | Including interviewee perceptions related to ‘what worked’ in implementing the work stream and why, and impact on interactive programme development | Systems  CFIR - process |
| X.4.8 Reflection and learning (negative) - iterative work stream development | Including interviewee perceptions related to ‘what didn't work’ in implementing the work stream and why, what they would do differently in future, and impact on interactive programme development | Systems  CFIR - process |
| X.4.9 Perceptions about reflection and learning | Perceptions about and instances of opportunities for reflection and learning in relation to the workstream to occur, utility of reflection, perception re whether reflection leads to change in practice | CFIR - process  NPT  Systems |

**Part C: Overarching JU:MP evaluation**

This section relates to the discrete evaluation work packages that JU:MP comprises - there is a separate theme for each evaluation work package. Data that is specific to an evaluation work package should be coded into the respective theme.

Theme 17: Process evaluation (strategic, neighbourhood, end user - qualitative) work package; Theme 18: Effectiveness evaluation (cohort, control trial, pilot trial - including mechanisms of change) work package; Theme 19: Organisation evaluation (organisational outcomes, minimum data-set) work package

| x.1 Research and evaluation characteristics | | |
| --- | --- | --- |
| X.1.1 Knowledge about research and evaluation characteristics | Knowledge about research / evaluation work package. This refers to anything the stakeholder perceives the research / evaluation to be ‘about’ - still code here if inaccurate perception. Include references to level of adaptability, complexity, quality, cost of the research. | TDF - knowledge  CFIR - knowledge about the intervention; design quality; costs; adaptability; complexity |
| X.1.2 Source of evaluation approach and strength of underpinning evidence | Perceptions of interviewees about how and by whom the research package has been developed, including for example: in line with implementation plan, theoretical basis, co-production; quality / strength of evidence underpinning research approaches (e.g. gold standard vs pragmatic approach).  **Exclude iterative development of research / evaluation work package based on** **process evaluation and learning (code to x.4.1 or x.4.2).** | CFIR - intervention source, quality and strength of evidence |
| X.1.3 Feelings about research / evaluation work package | Feelings about the research / evaluation work package including what the interviewee likes and doesn't like about the research / evaluation approach, perceptions about what ‘works well’ and why - about research design. **Exclude impact of research (code to x.3), reflection on implementation of research that leads to adapted practice (code to x.4.1 or x.4.2).** | Affective attitude (Sekhon)  CFIR - beliefs about the intervention, design quality, adaptability, complexity |
| X.1.4 Compatibility with the wider JU:MP (evaluation) approach | Detail re any connection, synergy or tension between the research and the values / principles of the overarching JU:MP programme, evaluation, or other research / evaluation components. **Exclude: tensions / compatibility related to implementation (code to x.2.3.3)** | CFIR - adaptability, complexity |
| X.2 Research / evaluation implementation | | |
| X.2.1 Research / evaluation work package implementation activity | Workstream strategic implementation activity - Engagement of implementers, including how researchers trained / engaged to deliver the research; carrying out / accomplishing implementation according to plan (what happened, when, how, by whom etc) | CFIR - implementation / process  NPT |
| X.2.2 Factors influencing implementation of the research / evaluation | | |
| X.2.2.1 Characteristics of ‘deliverers’ | Capability and motivation of researchers to deliver the research work package. Including, for example, knowledge of relevant topics, skills, relevant experience, values, how the work makes them feel, how emotional/health affect research delivery, managing workload | CFIR - characteristics of individuals  TDF - knowledge, skills, beliefs in capability, intention, identity, emotion, optimism |
| X.2.2.2 Characteristics of the research / evaluation work package | Characteristics of the research / evaluation work package that make it easier / harder to implement. | CFIR - intervention characteristics |
| X.2.2.3 Elements of the wider JU:MP programme | Characteristics of the wider JU:MP programme, including the wider research/evaluation, that makes the workstream easier / harder to implement. Including the influence of the funding organisation. | CFIR - inner setting |
| X.2.2.4 Core JU:MP team context | Factors related to the core JU:MP team that make it easier / harder to deliver the research / evaluation work package. Including: Social architecture, age, maturity, size etc. of the JU:MP team, ‘culture’ of the team (including whether feel valued as part of the team, safe to try new methods etc.).  The environmental context of the inner / core JU:MP team including networks and communication processes, the physical environment (office space etc.) access to resources, commitment of wider team to delivering research | TDF - social environment; environmental context and resources  CFIR - inner setting; structural characteristics; impl climate, relative priority, readiness for impl; networks and comms; available resources |
| X.2.2.5 Programme leadership | Programme leadership factors influencing delivery of the research, including: Commitment, involvement, and accountability of leaders and managers, including the extent to which leaders inspire / motivate the team; including team leads, AB board, JU:MP exec board. Incentives for deliverers such as goal-sharing awards, performance reviews / goal setting and feedback processes, promotions, and less tangible incentives such as increased stature or respect | CFIR - inner setting; leadership engagement; organisational incentives and rewards; goals and feedback |
| X.2.2.6 Factors related to research / evaluation participants | Including any insight into the experience of research participants. For example, whether they enjoyed it, or found it boring (acceptability), how easy it was, whether questionnaires were comprehensible (feasibility), engagement with the research. | Sekhon - acceptability  COM-B / TDF  CFIR - patient needs + resources |
| x.2.2.7 Factors related to wider stakeholders and organisations | Including barriers / facilitators to engaging with wider stakeholders as part of the research. The perceived feasibility and acceptability of wider stakeholders supporting delivery of the research. | Sekhon - acceptability  COM-B / TDF |
| X.2.2.8 Delivery neighbourhood context | Factors related to the nature / characteristics of JU:MP neighbourhoods, that influence delivery of the research, including:Structural characteristics (size), culture, environmental context and resources such as greenspace / local assets, demographics of residents, history, how ‘joined up’ local stakeholders are etc. | CFIR - structure, networks, culture, impl climate, available resources  CFIR - outer setting; need |
| X.2.2.9 Wider contextual factors | Factors related to the LDP area, wider Bradford area e.g. environment and resource, policies and local politics, and wider regional / (inter)national social, cultural, political, environmental factors that influence delivery of the research e.g. COVID-19 | CFIR - outer setting; cosmopolitanism, external policies, peer pressure |
| X.3 Research / evaluation impact | | |
| X.3.1 Beliefs about consequences | Expected / anticipated (not actual) consequences of JU:MP; the extent to which JU:MP is perceived as likely to achieve its purpose; positive / negative. | TDF - beliefs about conseq  Sekhon - perceived effectiveness |
| X.3.2 Impact of research activity | The influence of the research / evaluation activity, including, for example, on the attitudes or behaviours of key stakeholders, including the JU:MP team, steering groups, organisation, children and families. | Systems  Theory of Change  COM-B / TDF |
| X.3.3 Unintended negative consequences | Consequences of the research that were unintended and negative | Systems |
| X.4 Reflection and learning | | |
| X.4.1 Reflection and learning (positive) | Including interviewee perceptions related to ‘what worked’ in implementing the evaluation work package and why, and impact on interactive programme development | Systems  CFIR - process |
| X.4.2Reflection and learning (negative) - iterative work package development | Including interviewee perceptions related to ‘what didn't work’ in implementing the evaluation work package and why, what they would do differently in future, and impact on iterative work package development | Systems  CFIR - process |
| X.4.3 Perceptions about reflections and learning | Perceptions about and instances of opportunities for reflection and learning in relation to the evaluation work package to occur, utility of reflection, perception re whether reflection leads to change in practice | CFIR - process  NPT  Systems |
