## Additional file 6 for "A whole system approach to increasing children’s physical activity in a multi-ethnic UK city: a process evaluation protocol"

Observational framework

This summary sheet refers to any researcher observations completed as part of the process evaluation only. The purpose of this guidance is not to constrain data collection but rather to provide some common areas of initial and ongoing focus so that there is an agreed and overall structure for observational records. The researcher(s) will develop observational summaries and linked memos which relate to but also extend and add to the areas of focus identified below and on the observation record sheet.

Focus of observations

A researcher will observe selected and focused activity of JU:MP staff and stakeholders; namely, any workshops or steering groups that occur within neighbourhoods as part of developing and implementing JU:MP.

The purpose of the observations is to:

- Develop an understanding of the whole system and neighborhood contexts within which the JU:MP programme is taking place. Observations will be focused on physical activity and behaviour-change related information, including documenting regularities and irregularities of how organizations, teams, and individuals work and think in relation to physical activity.
- Describe and explain what JUMP staff and stakeholders are doing, including actions, decisions, how things were actually done vs plans, behaviour change techniques utilized
- Develop an understanding of changes that are occurring in the LDP area including tangible changes (e.g. changes to the environment) and intangible changes (e.g. how organisations talk about physical activity), and reflect on the changes by considering how they occurred, what the impact / consequences of the changes are and for whom, were they intended or unintended and positive or negative.
- Examine the extent to which the programme delivery and any identified system-changes, align with the programme theory (theory of change) and implementation plan.
- Identify barriers and facilitators to planning, delivering and implementing system-level and neighbourhood-level change as part of the JUMP programme, drawing on implementation theory constructs including: the extended normalization process theory and the consolidated framework for implementation research, and behavioral theory including capability opportunity motivation (COM-B) and the theoretical domains framework (TDF)
- Identify and develop intervention related topics or questions to explore in semi-structured interviews with relevant stakeholders.

Observation template

| **Name of project** | e.g. JU:MP |
| --- | --- |
| **Observation context** | e.g. which neighbourhood, workshop number, particular focus of the workshop |
| **Date and time of observation** | Include full length of session being observed, and the length of the observation, if these differ |
| **Observer name / role** | e.g. research fellow, intern, and whether usually work on JUMP evaluation |
| **A description of the session being observed** | |
| These are general observations, to capture a sense of what happened during session. No personal or identifiable information will be collected e.g. attendees, schools, organisations should be anonymised. This is a guide for the researcher indicating areas to capture in observations, drawn from Spradley (1980):   - Capture a sense of space of where the session took place (including built environment, how the activity was organised within space, how attendees used space) - Capture a sense of who is in attendance (and who may be missing) - Description of the content of the session including the organisation and structure of the session e.g. items discussed in meetings, workshop content including activities. If there is an agenda / plan available for the session, provide commentary on the extent to which the session follows / deviates from this - Description of interactions between people present including between deliverers / facilitators / chairs, between attendees, between deliverers / facilitators and attendees, and between the researcher and other attendees (where relevant to the session / research) - Description of engagement of attendees with session (for example, description of patterns of behaviour (e.g. body language, comments) suggesting engagement or disengagement, why etc. - Description of documents / materials utilised during the session including reference to where these are stored (if appropriate / relevant for documentary analysis) - Any problems/issues that occurred (including any issues raised by attendees) - Description of informal / unplanned activity that appears to have influenced the session | |
| **Reflective comments (memo)** | |
| These are reflective comments based on the researchers’ ideas, thoughts and feelings about the session. However, memos are not simply "ideas." They are involved in the formulation and revision of explanations for the processes observed and are utilized as part of the data analysis process. The memo should also reflect on how the observed session relates to theory underpinning the research and the research questions. This is a guide for researchers, indicating possible areas of reflection:   - Description of what went well /could have been improved in the session and why - Identification of behaviour change techniques utilised (intentionally or unintentionally) - Thoughts related to barriers and facilitators to planning, delivering and implementing change as part of the JU:MP programme drawing on implementation theory constructs and behavioural theory - Reflections on what system-level change has occurred, what led to these changes, what is the impact / consequences of the change and for whom, was this intended or unintended, what is visible and not visible, which voices have inputted into this learning, and has anything unexpected happened - Reflection on the extent to which the session, and any identified changes, align with the programme theory (theory of change), JU:MP framework, and implementation plan. - How observations link to NEF themes: place, audience, leadership, learning, partners, commitment, outcomes, sustainability ) | |
| **Possible points / issues to follow up in interviews or other future data collection** | |
| Please record here any identified areas that it may be useful to follow up on in future data collection that is not already recorded as part of the data collection plan. If possible note *who* to follow up with. | |

*Behaviour change techniques*
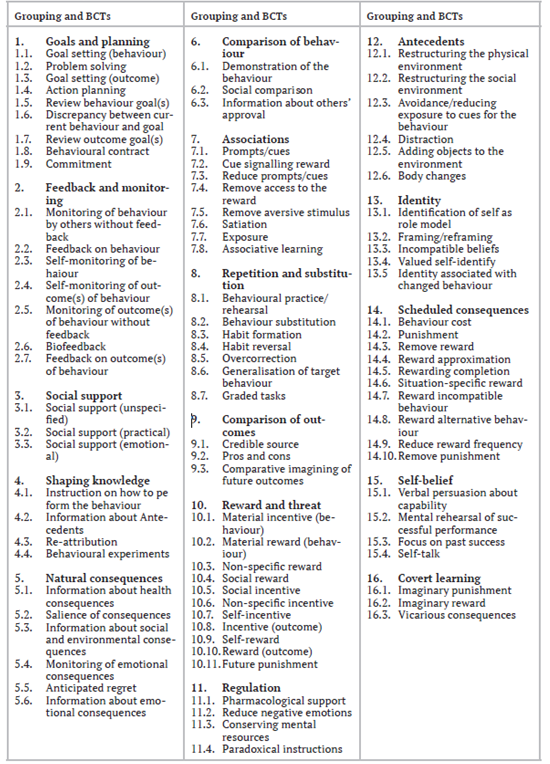

*COM-B - Michie et al., 2011*

COM-B stands for Capability Opportunity Motivation – Behaviour. The central tenet of the model is that for any behaviour to occur:

1. there must be the ‘capability’ to do it: the person or people concerned must have the physical strength, knowledge, skills, stamina etc. to perform the behaviour;
2. there must be the ‘opportunity’ for the behaviour to occur in terms of a conducive physical and social environment: e.g. it must be physically accessible, affordable, socially acceptable and there must be sufficient time;
3. there must be sufficient strong ‘motivation’: i.e. they must be more highly motivated to do the behaviour at the relevant time than not to do the behaviour, or to engage in a competing behaviour.

Each of these components can be divided heuristically into two types. Capability can be either ‘physical’ or ‘psychological’. Opportunity can be ‘physical’ (what the environment allows or facilitates in terms of time, triggers, resources, locations, physical barriers, etc.) or ‘social’ (including interpersonal influences, social cues and cultural norms). Motivation may be ‘reflective’ (involving self-conscious planning and evaluations (beliefs about what is good or bad) or ‘automatic’ (processes involving wants and needs, desires, impulses and reflex responses).
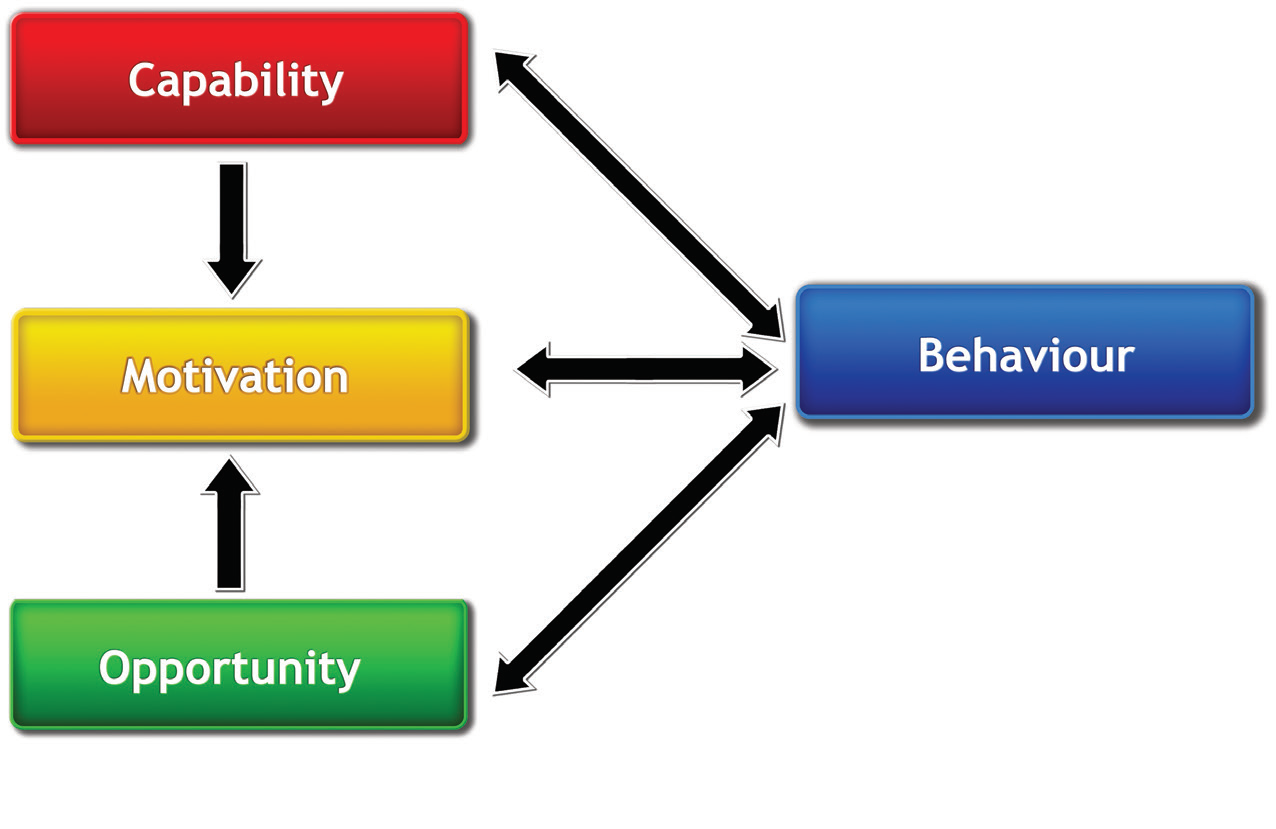

***Theoretical domains framework - Cane et al., 2012***

The TDF is an integrative framework synthesising key theoretical constructs used in relevant theories and was developed in a collaboration between psychologists and implementation researchers.

The framework consists of 14 domains: knowledge; skills; memory, attention and decision processes; behavioural regulation; social/professional role and identity; beliefs about capabilities; optimism; beliefs about consequences; intentions; goals; reinforcement; emotion; environmental context and resources; and social influences.

*Consolidated framework for implementation research*

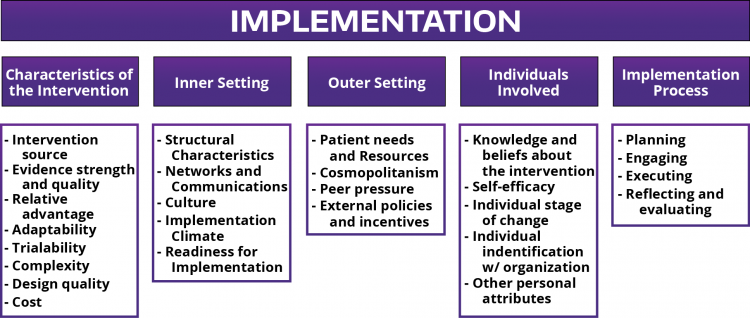

*Normalisation process theory*

NPT identifies factors that promote and inhibit the routine incorporation of complex interventions into everyday practice. It also explains how these interventions work, looking not only at early implementation, but beyond this to a point where an intervention becomes so embedded into routine practice that it is normalised. NPT characterises implementation as a social process of collective action. The social processes involve interactions between emergent expressions of agency, and dynamic elements of context. NPT consists of four constructs: coherence, cognitive participation, collective action, and reflexive monitoring.

(1) **Coherence** is the **sense-making work** that people do individually and collectively when they are faced with the problem of operationalizing some set of practices.
(2) **Cognitive Participation** is the **relational work** that people do to build and sustain a community of practice around a complex intervention.
(3) **Collective Action** is the **operational work** that people do to enact a set of practices, whether these represent a new technology or complex healthcare intervention. The components reflect qualities of complex interventions, rather than the character of the work that these involve.
(4) **Reflexive Monitoring** is the **appraisal work** that people do to assess and understand the ways that a new set of practices affect them and others around them.

*Guidance from the National Evaluation Framework*

The Local Delivery Pilot National Evaluation Partner (IFF) have provided some ‘thought starters’ linked to each of the themes that each LDP is required to report on, on a 6-monthly basis. These should be considered when writing the reflective comments.

| **Theme** | **Thought starters…** |
| --- | --- |
| **Place** | - How has your understanding of local challenges evolved over time? - How do your activities, efforts and investments respond to these challenges? - The effects of external factors that are outside the control of the place or pilot? |
| **Audience** | - How are you using local perspectives and information to make your approach locally relevant? - How are citizens and communities being engaged and empowered? - The extent to which the provision, delivery, services and investment going into communities is suited to the priorities of the target group/s and intended recipients? |
| **Leader- ship** | - Who is taking a leadership role? - How are local power dynamics evolving? - Have you seen any change in how decisions are made – who has a voice and influence? - How are problems identified and addressed? - How are leadership behaviours changing? |
| **Learning** | - How are you generating knowledge on what is and what is not working? - How are you using this knowledge and understanding to adapt and improve what you’re doing? - Where are the feedback loops that take information about what is happening and working on the ground to leaders and decision makers? |
| **Partners** | - Who is involved in the pilot activity? - How are connections and relationships evolving - what are their strengths and weaknesses? - How are partners showing understanding of and adapting to each others’ values and viewpoints? |
| **Commit-ment** | - How is capacity within the system changing – in terms of people, skills, time – and where it is directed? - How are key actors balancing wider responsibilities with the pilot? - What factors help to build and maintain momentum, and how? - What factors create distraction and block progress, and how? |
| **Outcomes** | - What value is being created? Who is this valuable to/for? - What are the main challenges and barriers to achieving what you set out to achieve? - Have you seen any unintended consequences arising? What are they, how have you responded to these, and why? |
| **Sustain- ability** | - Are you seeing any signs of sustainable elements or activities? - What will ensure that the changes you are making can be maintained (if they are working)? - What are the barriers to sustainability that you are observing? - What were the major factors which influenced the achievement or non-achievement of sustainability of the programme or project? |

*TDF Definitions*

| **Theme** | **Definition** | **Example** |
| --- | --- | --- |
| Knowledge | An awareness of the existence of something | Knowledge about physical activity interventions |
| Skills | An ability or proficiency acquired through practice | Ability to support physical activity interventions |
| Memory, attention and decision process | The ability to retain information, focus selectively on aspects of the environment and choose between two or more alternatives | Memory of past physical activity support sessions |
| Behavioural Regulation | Anything aimed at managing or changing objectively observed or measured actions | Reference to any behavioural systems that help track the physical activity interventions |
| Social/professional role and identity | A coherent set of behaviours and displayed personal qualities of an individual in a social or work setting | How the physical activity support interventions coincide with their professional identity |
| Beliefs about capabilities | Acceptance of the truth, reality or validity about an ability, talent or facility that a person can put to constructive use | How difficult they believe it is to carry out the physical activity |
| Optimism | The confidence that things will happen for the best or that desired goals will be attained | How confident they are that the physical activity support will have an impact |
| Beliefs about consequences | Acceptance of the truth, reality or validity about outcomes of a behaviour in a given situation | What consequences they believe will occur from the physical activity support |
| Intentions | A conscious decision to perform a behaviour or a resolve to act in a certain way | Any decisions they made for the physical activity interventions |
| Goals | Mental representations of outcomes or end states that an individual wants to achieve | Goals they want to achieve from the physical activity interventions |
| Reinforcement | Increasing the probability of a response by arranging a dependent relationship, or contingency, between the response and a given stimulus | Incentives they have to get people to engage in physical activity |
| Emotion | A complex reaction pattern, involving experiential, behavioural and physiological elements by which the individual attempts to deal with a personally significant matter or event | How physical activity makes them feel |
| Environmental context and resources | Any circumstance of a person’s situation or environment that discourages or encourages the development of skills and abilities, independence, social competence and adaptive behaviour | Any factors that affect physical activity being done |
| Social influences | Those interpersonal processes that can cause individuals to change their thoughts, feelings or behaviours | How social factors influence physical activity |
