## Additional file 5 for "A whole system approach to increasing children’s physical activity in a multi-ethnic UK city: a process evaluation protocol"

### NETWORK MAPPING

This survey is designed to understand relationships between different people and organisations within the neighbourhoods that JU:MP is working with. Below is a list of different stakeholders and organisations that work within this area. Please indicate for each:

1) Whether you were aware of them before today
2) How many times you have interacted or collaborated with them in the last 6 months
3) How many of these times were related to children's physical activity.

There is also space at the end to add additional stakeholders or organisations that work within this area, who you are aware of and/or have interacted or collaborated with.

| STAKEHOLDER OR ORGANISATION NAME | Were you aware of them before today? | How many times have you interacted or collaborated with them in the last 6 months? | How many of these times were related to children's physical activity? |
|---|---|---|---|
| | YES/NO | ADD NUMBER | ADD NUMBER |

We are family BORNINBRADFORD

JOIN US: MOVE. PLAY.

ACTIVE BRADFORD

SPORT ENGLAND

LOTTERY FUNDED
