## Supplementary figures and images for "A whole system approach to increasing children’s physical activity in a multi-ethnic UK city: a process evaluation protocol"

### Additional file 2

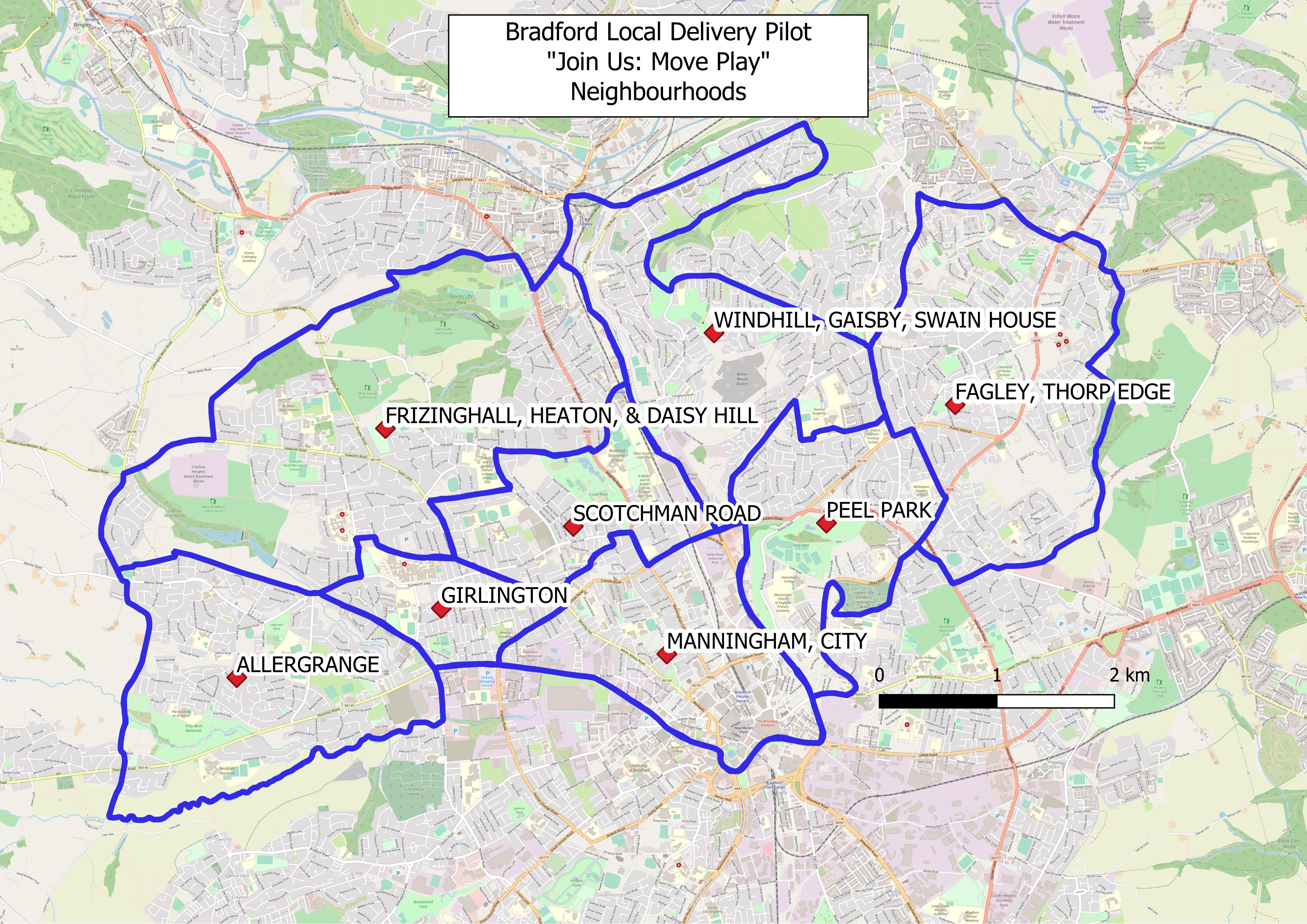
